# Inferring effects of mutations on SARS-CoV-2 transmission from genomic surveillance data

**DOI:** 10.1101/2021.12.31.21268591

**Authors:** Brian Lee, Ahmed Abdul Quadeer, Muhammad Saqib Sohail, Elizabeth Finney, Syed Faraz Ahmed, Matthew R. McKay, John P. Barton

## Abstract

New and more transmissible variants of SARS-CoV-2 have arisen multiple times over the course of the pandemic. Rapidly identifying mutations that affect transmission could improve our understanding of viral biology and highlight new variants that warrant further study. Here we develop a generic, analytical epidemiological model to infer the transmission effects of mutations from genomic surveillance data. Applying our model to SARS-CoV-2 data across many regions, we find multiple mutations that substantially affect the transmission rate, both within and outside the Spike protein. The mutations that we infer to have the largest effects on transmission are strongly supported by experimental evidence. Importantly, our model detects lineages with increased transmission even at low frequencies. As an example, we infer significant transmission advantages for the Alpha, Delta, and Omicron variants shortly after their appearances in regional data, when their local frequencies were only around 1-2%. Our model thus facilitates the rapid identification of variants and mutations that affect transmission from genomic surveillance data.

## Introduction

Viruses can acquire mutations that affect how efficiently they infect new hosts, for example by increasing viral load or escaping host immunity ^1–4^. The ability to rapidly identify mutations that increase transmission could inform outbreak control efforts and identify potential immune escape variants ^5–9^. However, estimating how individual mutations affect viral transmission is a challenging problem.

To address this challenge, we developed a method to infer the effects of single nucleotide variants (SNVs) on viral transmission that systematically integrates genomic data from different outbreak regions. Our analytical approach is based on a simple epidemiological model, allowing it to be efficiently applied to large data sets and opening the door to future theoretical extensions. Our method is also automatic in the sense that it relies only on sequence data and does not require, for example, clustering sequences into discrete “variants.” An additional advantage of our approach is that relative changes in viral transmission are statistically explained in terms of the specific mutations that different viruses bear, highlighting mutations that may be especially biologically important. For clarity, we refer to non-reference nucleotides (including deletions or insertions) as SNVs and viral lineages possessing common sets of SNVs as variants. Simulations show that our approach can reliably estimate transmission effects of SNVs even from limited data.

We applied our method to more than 7.4 million SARS-CoV-2 sequences from 149 geographical regions to reveal the effects of mutations on viral transmission throughout the pandemic. While the vast majority of SARS-CoV-2 mutations have negligible effects, we readily observe increased transmission for sets of SNVs in Spike and other hotspots throughout the genome.

Importantly, our approach is sensitive enough to identify variants with increased transmission before they reach high frequencies. This is demonstrated by studying the rise of the Alpha and Delta variants in Great Britain and Omicron in South Africa. We reliably infer increased transmission for these variants soon after their emergence, when their frequency in the region was only around 1-2%. An untargeted search for sets of mutations that strongly increase viral transmission also reveals multiple collections of SNVs belonging to well-known variants. Collectively, these data show that our model can be applied for the surveillance of evolving pathogens to robustly identify variants with transmission advantages and to highlight key mutations that may be driving changes in transmission.

## Results

### Epidemiological Model

To quantify the effects of mutations on viral transmission, we developed a generalized Galton-Watson-like stochastic branching process model of disease spread (Methods). Branching processes have been frequently used to model the stochastic numbers of infections in a population ^10,11^. Our model incorporates superspreading by drawing the number of secondary infections caused by an infected individual from a negative binomial distribution with mean *R*, referred to as the effective reproduction number, and dispersion parameter *k* (refs. ^12–17^). Multiple variants with different transmission rates are included by assigning a variant *a* an effective reproduction number *R*_*a*_ = *R*(1 + *w*_*a*_). Under an additive model, the net increase or decrease in transmission for a variant is the sum of the individual transmission effects *s*_*i*_ for each SNV *i* that the variant contains. In analogy with population genetics, we refer to the *w*_*a*_ and *s*_*i*_ as selection coefficients.

We then apply Bayesian inference to estimate the transmission effects of SNVs that best explain the observed evolutionary history of an outbreak. To simplify our analysis, we use a path integral technique from statistical physics, recently applied in the context of population genetics ^18^, to efficiently quantify the probability of the model parameters given the data (for details, see Supplementary Information). This allows us to derive an analytical estimate for the maximum *a posteriori* selection coefficients ***ŝ***, normalized per serial interval, for a given set of viral genomic surveillance data,

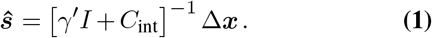

Here Δ***x*** is the change in the SNV frequency vector over time, *γ′* is a rescaled regularization term proportional to the precision of a Gaussian prior distribution for the selection coefficients *s*_*i*_ (Methods), and *I* is the identity matrix. The dispersion parameter *k* and number of infected individuals *N*, analogous to a population size in population genetics, are absorbed into the definition of *γ*^*′*^. *C*_int_ is the covariance matrix of SNV frequencies integrated over time, and accounts for competition between variants as well as the speed of growth for different viral lineages (Supplementary Information). Data from multiple outbreaks can be combined by summing contributions to the integrated covariance and frequency change from each individual trajectory (Methods). Our theoretical model could also be extended to incorporate additional features of disease transmission, such as the travel of infected individuals between different outbreak regions.

### Validation in simulations

To test our ability to reliably infer selection, we analyzed simulation data using a wide range of parameters. We found that inference is accurate even without abundant data, especially when we combine information from outbreaks in different regions (**Fig. 1, Supplementary Fig. 1**). Because we model the evolution of relative frequencies of different variants, accurate inference of selection does not require the knowledge of difficult-to-estimate parameters such as the current number of infected individuals or the effective reproduction number (Methods). Simulations also demonstrated that our model is robust to variations in effective reproduction numbers in different regions (**Supplementary Fig. 2**).

**Fig. 1.**
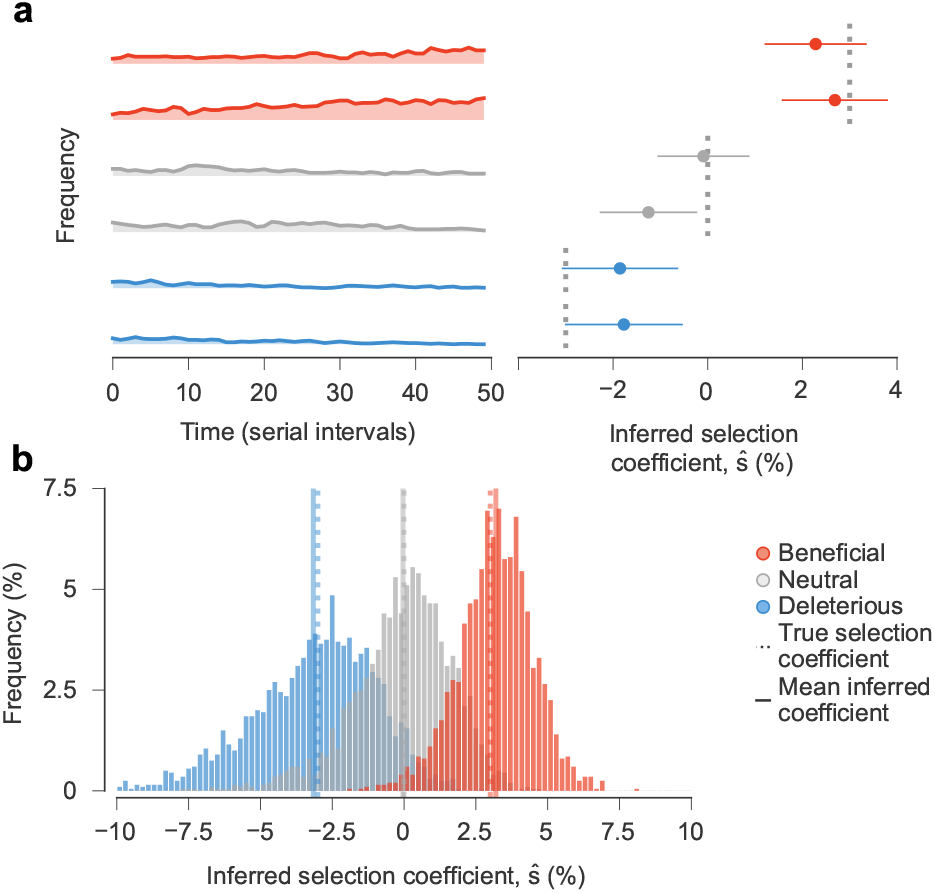
Our approach accurately estimates transmission effects of mutations in simulations. Simulated epidemiological dynamics beginning with a mixed population containing variants with beneficial, neutral, and deleterious mutations. **a**, Selection coefficients for individual SNVs, shown as mean values *±* one theoretical s.d., can be accurately inferred from stochastic dynamics in a typical simulation (Supplementary Information). **b**, Extensive tests on 1,000 replicate simulations with identical parameters show that inferred selection coefficients are centered around their true values. Deleterious coefficients are slightly more challenging to accurately infer due to their low frequencies in data. *Simulation parameters*. The initial population is a mixture of two variants with beneficial SNVs (*s* = 0.03), two with neutral SNVs (*s* = 0), and two with deleterious SNVs (*s* = −0.03). The number of newly infected individuals per serial interval rises rapidly from 6,000 to around 10,000 and stays nearly constant thereafter. Dispersion parameter *k* is fixed at 0.1.

### Global patterns of selection in SARS-CoV-2

We studied the evolutionary history of SARS-CoV-2 using genomic data from GISAID ^19^ as of January 26, 2024. We separated data by region and estimated selection coefficients jointly over all regions (Methods). After filtering regions with low or infrequent coverage, our analysis included more than 7.4 million SARS-CoV-2 sequences from 149 different regions, containing 1,398 nonsynonymous SNVs observed at nontrivial frequencies.

Our analysis reveal that, while the majority of SNVs were nearly neutral, a few dramatically increased viral transmission (**Fig. 2a**, **Supplementary Table 1**). We observe clusters of SNVs with strong effects on transmission along the SARS-CoV-2 genome (**Fig. 2b**). The highest density of SNVs that increase transmission is in Spike, especially in the S1 subunit (**Supplementary Fig. 3**). Of the top 20 mutations that we infer to be most strongly selected, 16 are in Spike (**Supplementary Table 1**). However, SNVs with a strong selective advantage are also found in other proteins, especially in N, NSP4, NSP6, and NSP12.

**Fig. 2.**
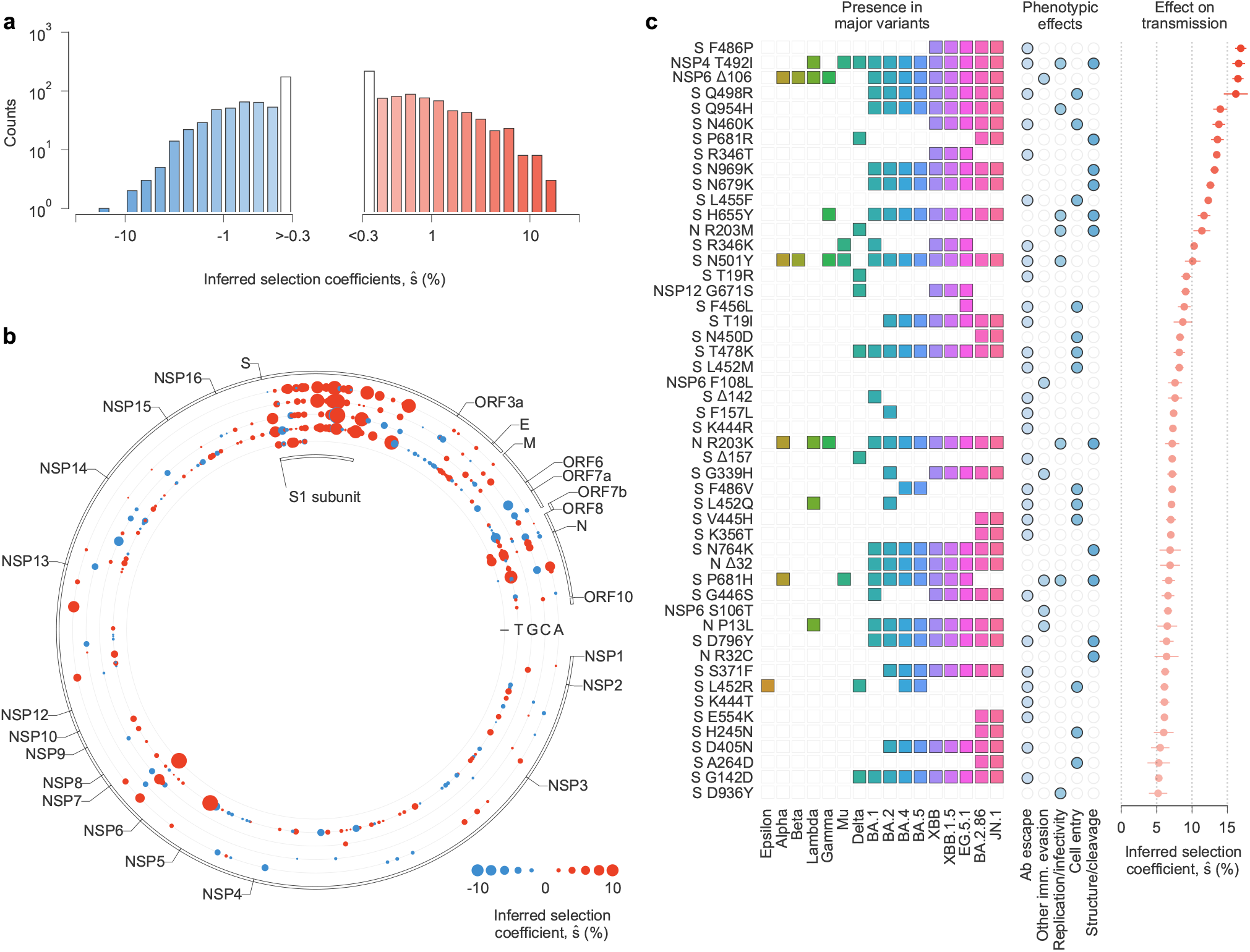
Inferred transmission effects of SARS-CoV-2 mutations. **a**, The majority of the 1,320 nonsynonymous SNVs included in our study are inferred to have negligible effects on transmission (that is, *ŝ* close to zero). However, a few SNVs have strong effects, as evidenced by a large value of *ŝ*. **b**, Patterns of selection across the SARS-CoV-2 genome. Beneficial SNVs often cluster together in the genome. Clustering is especially apparent for the S1 subunit of Spike, where many SNVs that are inferred to have the largest effects on transmission are located. **c**, Top 50 mutations inferred to increase SARS-CoV-2 transmission the most, the major variants in which these mutations are observed, their phenotypic effects, and selection coefficients (see **Supplementary Table 1**). The same colors are used to represent each major variant in **Figs. 3-4** and **Supplementary Fig. 4**. We cluster experimental phenotypic results into five categories: antibody evasion; other immune evasion; increases in replication and/or infectivity; ACE2 receptor binding and cell entry; and mutations affecting protein structure and/or cleavage.

### Mutations inferred to strongly increase transmission

The top 50 mutations inferred to increase SARS-CoV-2 transmission the most are listed in **Fig. 2c** and **Supplementary Table 1**. Experimental evidence directly or indirectly supports 48 of these 50 inferences. For clarity, we will reference mutations at the amino acid level rather than the underlying SNVs, which are also given in **Supplementary Table 1**.

Spike mutations F486P, Q498R, Q954H, N460K, P681R, R346T, N969K, and N679K comprise 8 of the top 10 mutations, and all have demonstrated functional effects that could increase transmission ^20–25^. Similarly, Spike mutations in the receptor binding motif (RBM) such as F486P, Q498R, N460K, N450D, T478K, N501Y, L452R, and the so-called FLip mutations L455F and F456L appear prominently in our analysis, comprising 9 of the top 25 mutations. Most of these mutations have been shown to increase resistance to RBM-specific neutralizing antibodies ^20–22,24,26^ and the majority also enhance ACE2 receptor binding ^4,21,27–31^.

Of these, N501Y (*ŝ* = 10.1%, ranked 15th) is shared by almost all major SARS-CoV-2 variants. Q498R, N460K, and T478K are shared by all Omicron variants. Beyond the functional effects above, N501Y is known to increase transmission of infection ^32^ and to help maintain Spike in an active conformation for receptor recognition ^21^. Eight Spike N-terminal domain (NTD) mutations/deletions (T19R/I, Δ142, Δ157/F157L, H245N, A264D, and G142D) are also strongly selected. These lie in the antigenic supersite where mutations have been shown to decrease the neutralization potency of NTD-specific monoclonal antibodies ^21,33^. Spike mutations unique to the recently emerged Omicron variants BA.2.86 and JN.1 (N450D, V445H, K356T, E554K, H245N, and A246D) along with those found in the KP.2 and KP.3 variants which have become globally dominant in 2024 (R346T, F456L), rank among the top mutations identified in our analysis. All these mutations are known to impact either ACE2 receptor binding or antibody neutralization ^26,34–37^.

Research on viral transmission has naturally focused on Spike because of its role in viral entry and as a target of neutralizing antibodies. However, our analysis also reveals strongly selected mutations outside of Spike. These include the NSP4 mutation T492I, and Nucleocapsid mutations R203M/K, Δ32/R32C, and P13L. NSP4 mutation T492I (*ŝ*= 16.6%, ranked 2nd) was reported to increase viral replication and infectivity, enhance cleavage of the viral protease NSP5, and contribute to immune evasion based on experiments and animal models ^38^. Nucleocapsid mutation R203M (*ŝ* = 11.4%, ranked 13th) is in the linker region of the protein and enhances viral RNA replication, delivery, and packaging, which may increase transmission ^39^. Studies suggest that NSP6 mutations Δ106 and S106T (ranked 3rd and 38th, *ŝ*= 16.5 and *ŝ*= 6.6) and F108L (ranked 23rd, *ŝ*= 7.6) may increase transmission by interferon antagonism ^40^. We also find additional mutations outside of Spike, such as G671S in the RNA-dependent RNA polymerase NSP12 and Δ32 in N, that are highly selected and may be good targets for further experimental study. Our model thus highlights non-Spike mutations that may confer a selective advantage to emerging variants.

### Estimates of selection for major SARS-CoV-2 variants

We estimated the net increase in viral transmission relative to the WIV04 reference sequence for well-known SARS-CoV-2 variants by adding contributions from the individual variant-defining SNVs (**Fig. 3** and **Supplementary Fig. 4**, see Methods). Because our model uses global data and infers the transmission effects of individual SNVs, variants can be compared to one another directly even if they arose on different genetic backgrounds, or if they appeared in different regions or at different times. This also allows us to infer substantially increased transmission for variants such as Gamma or Mu, which never achieved the level of global dominance exhibited by variants like Alpha, Delta, Omicron, or XBB (**Supplementary Fig. 4**).

**Fig. 3.**
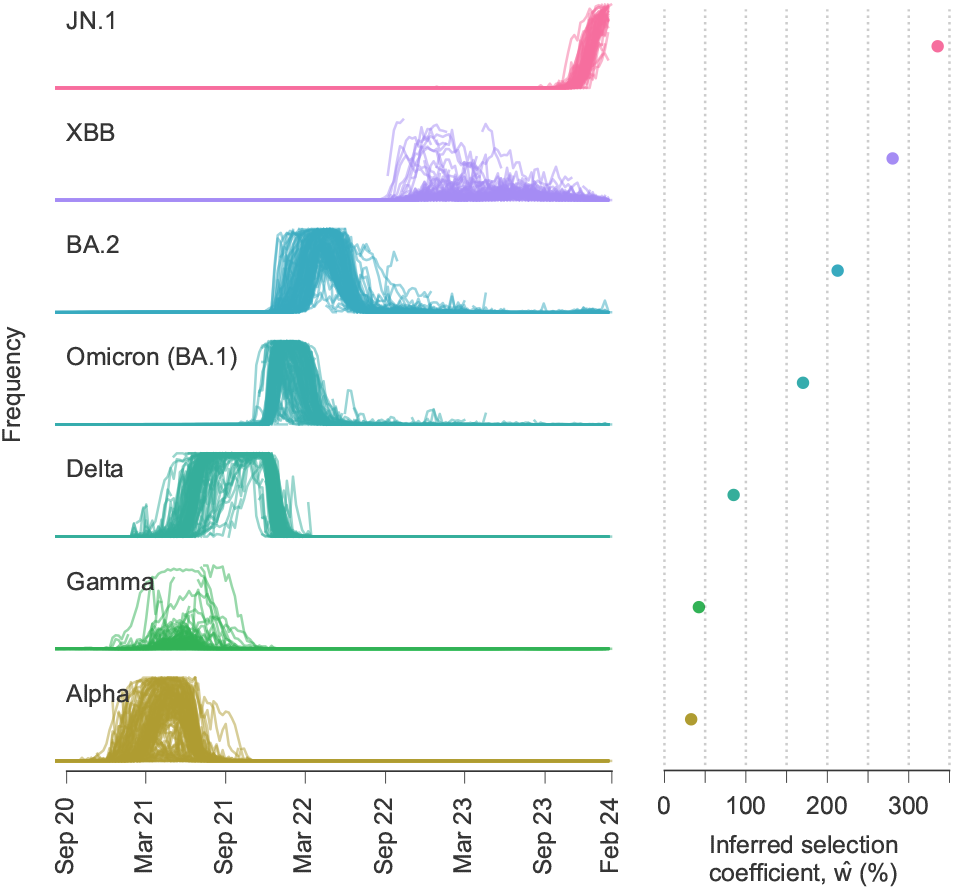
Multiple SARS-CoV-2 variants strongly increase transmission rate. Frequencies of major variants and their total inferred selection coefficients, shown as mean values *±* one s.d. from bootstrap subsampling of regional data (Methods), defined relative to the WIV04 reference sequence. Selection coefficients for variants with multiple SNVs are obtained by summing the effects of all variant-defining SNVs.

Our findings are consistent with past estimates that have shown a substantial transmission advantage first for Alpha and then for Delta relative to other pre-Omicron lineages ^41–43^. However, past estimates have varied substantially depending on the data source and method of inference. In different analyses, Delta has been inferred to have an advantage of between 34% and 97% relative to other pre-Omicron lineages ^41,42,44^. Similarly, Alpha has been estimated to increase transmission by 29% to 90% relative to pre-existing lineages in different regions ^5,41,45–47^. One advantage of our approach is that it can infer selection coefficients that best explain the growth or decline of variants across many regions, allowing for more even comparisons.

Over the period of data that we analyzed, Omicron and its subvariants display clear, large increases in transmission over past variants (**Fig. 3**). The transmission advantage of BA.1 (*ŵ*= 170%), which we estimate to be the least transmissible of Omicron subvariants, is roughly twice as large as the inferred selection coefficient for Delta (*ŵ*= 85%). More recent variants of Omicron, such as XBB (*ŵ*= 280%) are inferred to be substantially more transmissible.

In general, we find that the contributions of individual SNVs to the overall selection coefficient *ŵ*for a variant are very heterogeneous. A small fraction of mutations are responsible for most of the increase in transmission. As an example, **Supplementary Fig. 5** shows the relative contribution of each Alpha, Delta, and Omicron (BA.1) SNV to the total selection coefficient *ŵ*for the variant. In each case, fewer than 20% of SNVs are responsible for more than 80% of the increase in transmission.

### Detection of selection at low frequencies

We asked whether strong selection could be inferred for beneficial SNVs when they are still at low frequencies, before they dominate the viral population. To explore this, we considered the rise of the three major variants of concern (VOCs): Alpha and Delta in Great Britain, and Omicron (BA.1) in South Africa. We computed the inferred selection coefficient *ŵ*for each variant in each region at different points in time, as the VOCs began increasing in frequency. Selection coefficients were computed at different times by filtering the sequence data from GISAID to exclude sequences after a specific cutoff date. Note that this approach is different from previous sections, which used all data through January 26, 2024 to compute selection coefficients. To focus on selection for novel SNVs, we removed putative beneficial SNVs that had been previously observed in other VOCs from the estimates of *ŵ*.

We found that the inferred selection coefficients for novel Alpha SNVs rose rapidly as the variant was emerging (**Fig. 4a**). At the time that Public Health England labeled Alpha a variant of interest (VOI) ^48^, the inferred selection coefficient for novel Alpha SNVs was around 15%. When Alpha was declared a VOC ^49^, this had grown to around 45%. These statistics would indicate a substantial transmission advantage for Alpha relative to co-circulating variants. Notably, we inferred novel Alpha SNVs to be strongly beneficial even while the variant remained at low frequencies in Great Britain.

**Fig. 4.**
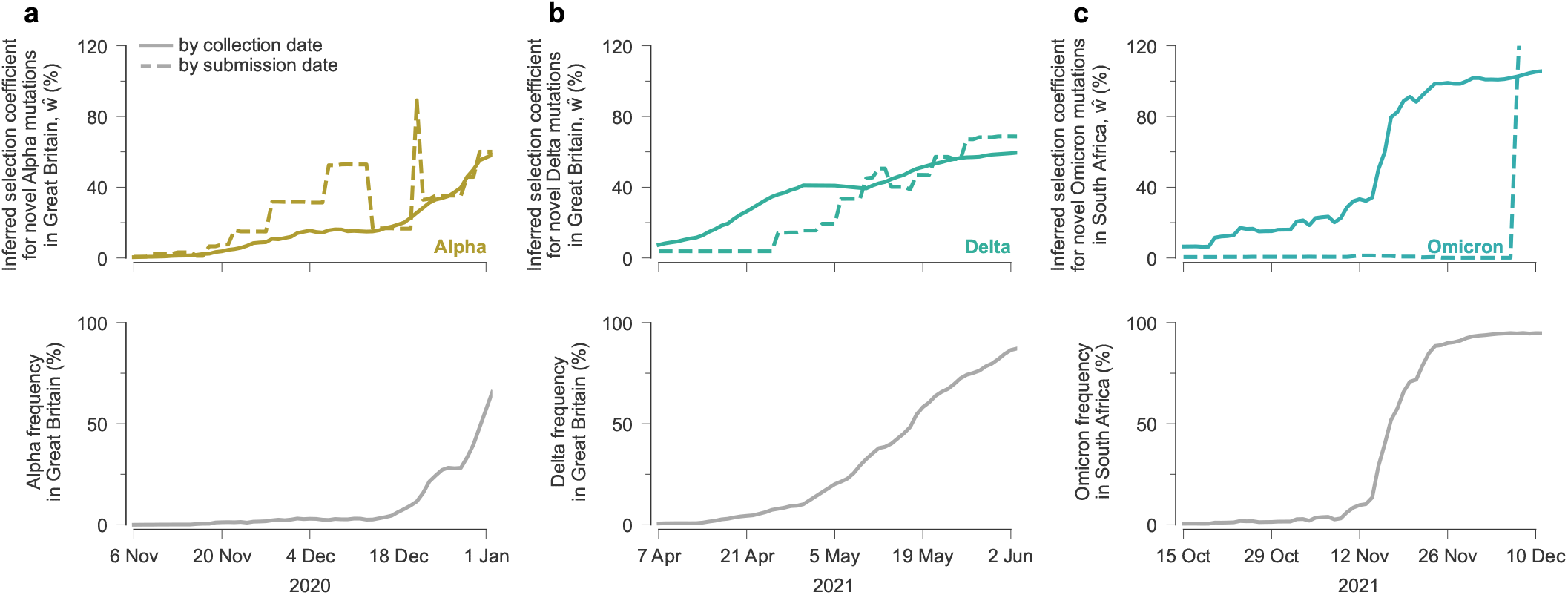
Our model rapidly infers increased transmission for Alpha, Delta, and Omicron (BA.1) SNVs. Inferred selection coefficients for novel SNVs in Alpha in Great Britain (**a**), Delta in Great Britain (**b**), and Omicron (BA.1) in South Africa (**c**) over time. Selection coefficients were computed over time according to GISAID data filtered by collection date or submission date. Selection coefficients given for a particular date include only data collected or submitted on or before that date. Variant frequencies are computed using sequence data filtered by collection date.

Similar analyses show that our model rapidly infers increased transmission for novel SNVs in Delta and Omicron. The selection coefficient for novel Delta SNVs in Great Britain was around 60-70% when it was classified as a VOC ^50^ (**Fig. 4b**). No full-length Omicron sequences were available on GISAID when it was designated as a VOC ^51^. However, the first Omicron data from South Africa uploaded on December 7, 2021, clearly revealed an enormous transmission advantage for Omicron (**Fig. 4c**).

In each of these examples, strong selection was detectable even for variants at low frequencies. To illustrate this point, we filtered SARS-CoV-2 sequence data by its collection date in each of these regions and computed the frequency of the Alpha, Delta, and Omicron variants over time. At the time that each variant reached a frequency of 2% in the population, their inferred selection coefficients for novel SNVs were 11%, 16%, and 21% for Alpha, Delta, and Omicron, respectively. These results show that our model can identify SNVs associated with higher transmission even when they are present in a small fraction of all infections in a region.

### Robust identification of beneficial SNVs

Identifying variants that increase transmission in real time could inform public health efforts and highlight important aspects of viral biology. However, the inherent stochasticity of infection and data collection makes accurate inferences difficult. For example, neutral or modestly deleterious SNVs may initially appear to be beneficial due to a transient rise in frequency despite having no selective advantage.

To explore the effects of fluctuations on estimates of selection, we first quantified the inferred selective advantage for all variants *ŵ*(including both SNVs and collections of SNVs that are strongly linked to one another, see Methods) in each region, for each day that data was submitted to GISAID. As in the previous section, data was filtered by submission date, such that selection coefficients computed for a specific date used only sequences that were submitted to GISAID on or before that date. Here, we progressively step through time in each region, adding sequences according to their submission date and re-analyzing the data in each region separately.

Although variation in sampling could produce temporary spikes in inferred selection coefficients, we reasoned that large *ŵ*are much more likely to be observed for variants with real, substantial advantages in transmission. To test this reasoning, we used the *ŵ*to identify variants with especially large inferred effects on transmission, which we refer to as high growth (HG) variants (*ŵ> θ* for some threshold value *θ*). In each region, we began at the first time point that data was submitted to GISAID and stepped through each subsequent upload date. At each step, we classified strongly linked SNVs with *ŵ> θ* as HG and excluded these SNVs from future analysis in the same region.

While it is difficult to conclusively determine whether the classification of a group of SNVs as HG is “correct” or “incorrect”, we conservatively assumed that (groups of) SNVs in major variants denoted by Greek characters or the B.1 variant should be correctly labeled as HG (true positives), and any other SNVs classified as HG constitute false positives. With this convention, the fraction of true positives increases steadily along with the threshold *θ*, such that more than 95% of variants classified as HG are true positives for *θ*≥18.5% (**Supplementary Fig. 6**). Thus, variants with inferred selection coefficients *ŵ>* 18.5% in any region and at any time are highly likely to have a substantial transmission advantage. This threshold could then be used to highlight new variants of particular interest.

We further studied the cumulative fraction of variant-defining SNVs that were classified as HG for 10 major SARS-CoV-2 variants, over time and in 7 broad geographical regions (**Supplementary Fig. 7**). Despite our stringent threshold of *θ* = 18.5%, a large fraction of variant-defining SNVs are ultimately found in HG groups in one or more regions. HG groups encompassing most SARS-CoV-2 variants were also independently detected across different regions, usually within a short period. Importantly, for these variants, around 10-30% of variant-defining SNVs were classified as HG *before* the variants began wide circulation among humans. This means that not only were some variant-defining SNVs observed in prior variants, they were also highlighted in our approach as SNVs that were likely to substantially increase SARS-CoV-2 transmission.

### Features of HG SNVs not in major variants

At the threshold value of *θ* = 18.5%, we found 38 groups of strongly linked SNVs that did not belong to major, Greek letter variants or B.1. Some of these groups of SNVs may have been identified as HG due to sampling noise. However, others may have biological effects that affect transmission, but not enough to outcompete more transmissible variants. Thus, we investigated whether SNVs in this list could have plausibly affected transmission.

Of the 38 groups, 12 sets of SNVs included Spike mutations with experimentally demonstrated effects or that lie in functionally important locations. Mutations A879S and A626S were experimentally shown to reduce sensitivity to convalescent sera ^52,53^. D138Y and W152R/L were shown to escape neutralization by specific antibodies ^54,55^, and N439K had reduced sensitivity to sera and antibodies ^52,56^. N439K and A520S increase binding to the ACE2 receptor ^56,57^. In addition, I794N lies on the fusion peptide and on the surface of the spike protein ^58^, while Q677P and S680P lie on the furin cleavage site ^59,60^. In summary, a substantial fraction of HG Spike SNVs that are not present in major variants could plausibly affect transmission, even if their effects are more modest than some SNVs in major variants.

## Discussion

Quantifying the effects of mutations on viral transmission is an important but challenging problem. We developed a flexible, branching process-based epidemiological model that provides analytical estimates for the transmission effects of SNVs from genomic surveillance data. Applying our model to SARS-CoV-2 data, we identified SNVs that substantially increase viral transmission, including both experimentally-validated Spike mutations and other, less-studied mutations that may be promising targets for future investigation. Importantly, we found that our model is sensitive enough to detect substantial transmission advantages for SNVs belonging to major variants even when they comprised only a small fraction of the total number of infections in a region.

Distinct from our method, current approaches to estimate changes in viral transmission often rely on phylogenetic analyses or fitting changes in variant frequencies to logistic or multinomial growth models ^5,46,47,61–63^. Phylogenetic analyses for viruses can be challenging due to a high degree of sequence similarity, which implies that the data can be explained equally well by a number of different trees ^64^, and they also typically rely on Markov chain Monte Carlo sampling that becomes intractable for large data sets. Growth models have been commonly applied to predict relative growth of SARS-CoV-2 variants, and have been incorporated into the popular NextStrain tool ^65^. These models can estimate the difference in transmissibility between one variant and others circulating in the same region. However, their estimates may be difficult to compare for variants that arose in other regions or with different genetic backgrounds, and they typically do not identify specific mutations responsible for changes in transmission.

Our approach differs from these due to our focus on explaining transmission differences between variants by the fitness contributions of individual SNVs. The scalable, analytical form of our estimator for fitness effects also allows for the natural integration of data from multiple regions. The predictions of our model are strongly supported by biological and experimental data. Phenotypic effects have been established for nearly all (i.e., 48 of the top 50; **Supplementary Table 1**) of the SNVs that we infer to be most beneficial for SARS-CoV-2 transmission. Our approach is based on a branching process epidemiological model of viral transmission. This is distinct from “black box” deep learning methods (including large language models) that have been proposed to address related but distinct problems, such as characterizing antigenic evolution and antibody escape dynamics ^66,67^.

The epidemiological model that we have introduced has limitations. We assumed a fairly short generation time, which is appropriate for a virus such as SARS-CoV-2. A different approach would be needed to consider the spread of viruses where many transmission events are from long-term infections, such as HIV. We also assume that SNVs contribute additively to fitness and that selection coefficients are constant in time. Our model does not delineate intrinsic (e.g., functional) effects of SNVs on transmission from selection advantages due to immune escape; though, for many of the SNVs inferred most strongly to affect transmission, there is independent experimental evidence to suggest that each (or both) of these factors are important (**Supplementary Table 1**). In principle, selection for immune escape is likely to be time-varying, as the buildup of population immunity reduces the selective advantage of escape mutations over time ^68^. Simulations show that if selection is time-varying, the constant selection coefficients that we infer reflect averages of time-varying selection over the time that the variant was observed (**Supplementary Fig. 8**). Epistasis could also lead to over- or under-estimation of selection coefficients for specific SNVs, but total contributions to transmission from multiple SNVs are typically estimated accurately (**Supplementary Fig. 9**). We have also assumed that serial intervals are constant in time, but variants may differ in the typical time between infections ^69^ which could influence relative growth rates. Differences in antigenicity could also generate fitness differences that are intransitive and which depend on immune history. A model that explicitly incorporates antigenicity would be needed to account for this effect. Finally, we note that no model based solely on dynamics, including ours, could distinguish the independent effects of different SNVs that exclusively appear together on the same genetic background.

Our ability to rapidly identify new, high growth variants is naturally limited by the public availability of sequence data. Time lags between when sequencing is performed and when sequences are uploaded, in particular, can lead to delays. As shown in **Fig. 4**, filtering sequences by collection date rather than submission date typically leads to much faster detection of variant growth. The disparity is especially large for Omicron: sequence data collected by mid-October 2021 already shows a substantial transmission advantage for this variant. In Great Britain, early Alpha sequences were significantly more likely to have short delays between collection and submission, causing Alpha sequences to be over-represented in early data and closing the gap between selection estimates. Even in this unusual case, however, earlier reporting substantially reduces noise. Thus, reducing the time between when sequencing is performed and when sequence data is publicly shared could facilitate the detection of new variants with increased transmission and help prepare for growing outbreaks. Our focus on quantifying the effects of individual mutations on viral transmission also mitigates some data limitations. Even in cases where sequence data for a novel variant is limited, emerging variants could be identified for further attention based on the presence of previously-observed mutations. For example, Alpha, Delta, and Omicron (BA.1) would have had estimated selection coefficients of *ŵ*= 18%, 17%, and 66%, respectively (relative to the WIV04 reference sequence), immediately prior to their first observations in sequence data. More generally, as shown in **Supplementary Fig. 7**, for multiple major variants there is evidence that some of their variant-defining SNVs substantially increase transmission prior to the wide circulation of those variants among humans.

While our study has focused on SARS-CoV-2, the epidemiological model that we have developed is very general. The same methodology could be applied to study the transmission of other pathogens such as influenza. Combined with thorough genomic surveillance data, our model provides a powerful method for rapidly identifying more transmissible viral lineages and quantifying the contributions of individual mutations to changes in transmission.

## Data Availability

Sequence data and metadata used in this study are available from GISAID. Processed data and code are available in the linked GitHub repository.

https://github.com/bartonlab/paper-SARS-CoV-2-inference

## ACKNOWLEDGEMENTS

We gratefully acknowledge all data contributors, i.e., the Authors and their Originating laboratories responsible for obtaining the specimens, and their Submitting laboratories for generating the genetic sequence and metadata and sharing via the GISAID Initiative, on which this research is based. The work of B.L., E.F., and J.P.B. reported in this publication was supported by the National Institute of General Medical Sciences of the National Institutes of Health under Award Number R35GM138233. The work of A.A.Q., M.S.S., and M.R.M. was supported by the Research Grants Council of the Hong Kong Special Administrative Region, China under Project No. T11-705/21-N; A.A.Q. and M.S.S. were also supported under Project No. 16204121. M.R.M. is the recipient of an Australian Research Council Future Fellowship (Project No. FT200100928) funded by the Australian Government.

## AUTHOR CONTRIBUTIONS

All authors contributed to methods development, data analysis, interpretation of results, and writing the paper. B.L. and J.P.B. led theoretical analyses. M.S.S. and B.L. led simulations. A.A.Q. led validation of SARS-CoV-2 inference results. J.P.B. conceptualized the project. J.P.B. and M.R.M. supervised the overall project.

## Methods

### Epidemiological model

We use a discrete time branching process to model the spread of infection. Individuals can be infected by any one of *M* viral variants, which are represented by genetic sequences ***g*** = {*g*_1_, *g*_2_, …, *g*_*L*_} of length *L*. For simplicity, we will first assume that alleles at each site *i* in the genetic sequence for variant *a* are either equal to the “wild-type” or reference 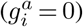 or mutants 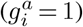. Later we will relax this assumption to consider genetic sequences with 5 possible states at each site (4 nucleotides or a gap). We call *n*_*a*_(*t*_*m*_) the number of individuals infected by variant *a* at time *t*_*m*_. To account for super-spreading, the number of newly infected individuals at time *t*_*m*+1_ follows a negative binomial distribution ^70–75^, *P* (*n*_*a*_(*t*_*m*+1_) |*n*_*a*_(*t*_*m*_), *k, R*_*a*_) = *P*_*NB*_ (*r, p*), where *r* = *n*_*a*_*k, p* = *k/*(*k* + *R*_*a*_), and *R*_*a*_ = *R*(1 + *w*_*a*_). Here *r* and *p* are the negative binomial distribution parameters, *k* is the dispersion, *R* is the effective reproductive number of the reference variant, and *w*_*a*_ encodes the variant dependence of the infectivity. The parameters *n, k*, and *R* can be time-varying. For instance, a time-varying *R* represents change in the number of susceptible and recovered individuals as well as the effects of public health interventions or changes in behavior that modify viral transmission.

Defining the frequency of variant *a* as *y*_*a*_ = *n*_*a*_*/*Σ_*b*_ *n*_*b*_, the probability that the frequency vector is ***y***(*t*_*m*+1_) = {*y*_1_(*t*_*m*+1_), *y*_2_(*t*_*m*+1_), …} given the initial frequency vector ***y***(*t*_0_), is

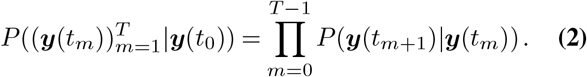

### Derivation of the estimator

Because (2) is difficult to work with directly, we follow the approach of ref. ^76^. We introduce a “diffusion approximation” where we assume that the total number of infected individuals is large and the effects of mutations on transmission are small. Similar approximations have been widely used in population genetics ^77–79^. Under these assumptions, the probability distribution for the variant frequencies satisfies a Fokker-Planck equation with terms derived from the first and second moments of the frequency changes *y*_*a*_(*t*_*m*+1_)− *y*_*a*_(*t*_*m*_) under the negative binomial distributions above.

However, the genotype space is high-dimensional (dimension 2^*L*^, with either a mutant or wild-type allele at each site) and undersampled, making inference of selection for genotypes extremely challenging. To simplify the inference problem, we assume that selection is additive, so the total selection coefficient *w*_*a*_ for a variant *a* is the sum of selection coefficients *s*_*i*_ for mutant alleles at each site *i*:

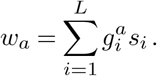

We can then derive a Fokker-Planck expression for the dynamics of mutant allele frequencies

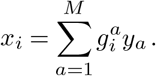

At the allele level, the Fokker-Planck equation has a drift vector given by

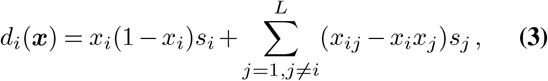

and a diffusion matrix

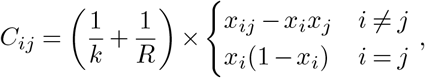

where *x*_*ij*_ is the frequency of infected individuals that have mutant alleles at both site *i* and site *j* at time *t*. In deriving (3) we have assumed that the selection coefficients satisfy *s*_*i*_ « 1 such that *w*_*a*_ « 1. Despite this technical assumption, our simulations demonstrate that selection can be accurately inferred even when selection is strong (**Supplementary Fig. 10**).

The drift vector describes the expected change in allele frequencies over time. Eq. (3) consists of two terms. The first describes the expected change in the frequency of allele *i* due to selection at that site. The second term accounts for linkage, that is, it quantifies how the genetic background alters the expected frequency change of an allele.

The Fokker-Planck equation can then be used to derive a path integral, which gives the probability of an entire evolutionary history or “path” (i.e., frequencies of genetic variants over time, 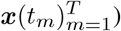). In Supplementary Information, we derive the path integral expression following a similar approach to the one described in ref. ^76^. The path integral is

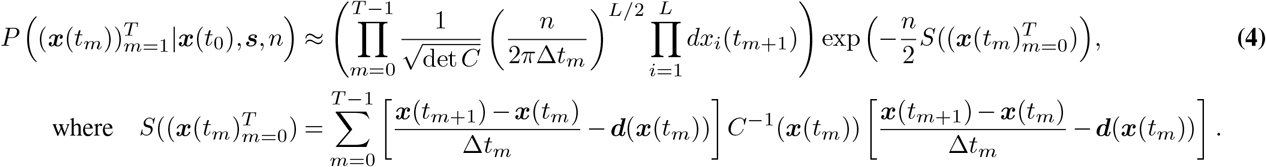

Here 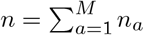 is the total number of individuals infected by all variants and Δ*t*_*m*_ = *t*_*m*+1_ − *t*_*m*_. The path integral in (4) has a form that is similar to the one obtained in ref. ^76^. The path integral quantifies the probability density for paths of mutant allele frequencies in the evolutionary history of the pathogen. We can then use Bayesian inference to find the maximum *a posteriori* estimate for the selection coefficients given the frequencies, the infected population size, the parameters *R* and *k*. The posterior probability of the selection coefficients is

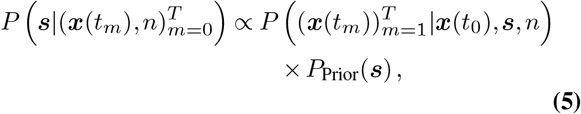

where 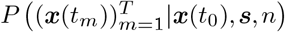 is the probability of a path given by (4) and the *P*_Prior_(***s***) is a Gaussian prior probability for the selection coefficients with zero mean and covariance matrix *σ*^2^*I*. Here, *I* is the identity matrix and *σ*^2^ is the variance of the prior. We call the precision *γ* = 1*/σ*^2^. In Supplementary Information we show that the selection coefficients that maximize (5) are

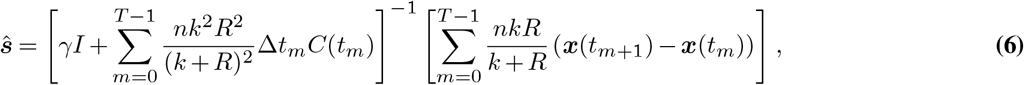

where the parameters *k, R*, and *n* are implicitly functions of *t*.

There are two interesting limiting forms of the estimator. First, we define the new matrix 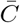 whose entries are

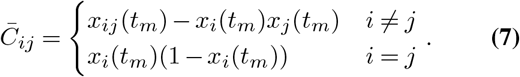

In the limit that *k* → ∞, the negative binomial distribution for new infections becomes a Poisson distribution with rate *λ* = *R*. In this special case, the model is equivalent to the Wright-Fisher model from population genetics. The estimator reduces to

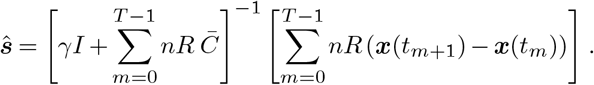

The opposite limit *k*→0 corresponds to a distribution for new infections with extremely heavy tails, i.e., one where super-spreading is dominant. In this case the drift in (3), which quantifies expected frequency changes due to selection, is unchanged. However, the diffusion matrix, which encodes linkage as well as the changes in frequency that are due to the stochastic nature of infection transmission, diverges. In this case, diffusion dominates the process entirely.

### Simplifying the estimator and robustness to incomplete knowledge of time-varying parameters

While our model has the ability to account for the time dependence of parameters appearing in (6), such as the infected population size *n*, the dispersion *k*, and the mean reproductive number *R*, these can be challenging to reliably estimate from data. However, we generally do not require full knowledge of these time-dependent parameters to accurately estimate selection.

In fact, due to finite sampling noise, estimates of selection produced by assuming constant (and incorrect) parameters are more accurate than estimates that use the true time-varying parameters (**Supplementary Fig. 11**). The naive estimator in (6) implies that time points or regions with larger *R, n*, or *k* should be weighted more heavily in the estimate. However, frequency information is always inaccurate due to noise from finite sampling, so weighing some time points or regions significantly more than others based upon the parameters alone means that undue weight is given to the uncertain information available from these times and regions.

For this reason, we assume parameters that are spatially and temporally constant in all of the following analysis as discussed below. This allows the estimator to be simplified substantially. If we assume constant parameters and scale the regularization *γ* by *nkR/*(*k* + *R*) in the numerator in (6), the parameter dependence in the numerator and the denominator is identical and cancels out (due to the additional factor of (*k* + *R*)*/kR* in the definition of the covariance matrix). With the same definition of the matrix 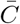 as above, and additionally defining 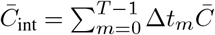 and *γ*^*′*^= *γnkR/*(*k* + *R*), the simplified estimator is given by

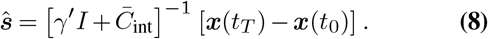

This form of the estimator is similar to the estimator for selection coefficients in the Wright-Fisher model ^76^, except that it omits contributions from the mutation term, because the mutation rate for SARS-CoV-2 is small. Practically, (8) has significant advantages over (6). The most important is that the difficult-to-estimate parameters *k* and *n* are no longer required. In addition, *R* does not need to be estimated. For methods of inferring these parameters as well as discussions about the difficulty of inferring them, see refs. ^80–89^.

### Extension to multiple regions and multiple SNVs at each site

The model can easily account for outbreaks in multiple regions or outbreaks at different times. If the probability of the evolutionary path in each region is independent, which is the case if there is no travel between regions, then the probability of all of the evolutionary paths in all of the regions is simply the product of the probabilities of the paths in each region, given by (4). Bayesian inference can be applied in the same way as before, resulting in the estimator

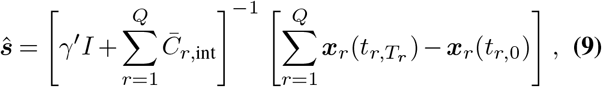

where *Q* is the number of regions, *t*_*r*_ is the time in region 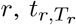 is the final time in region *r, t*_*r*,0_ is the initial time in region *r*, ***x***_*r*_ is the frequency in region *r*, and 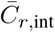 is the scaled integrated covariance matrix in region *r* given by integrating (7) over time. The estimator can further be extended to allow for multiple different nucleotides at each site by simply letting each different nucleotide have its own entry in the frequency vector *x*_*i*_. If there are *J* mutations at each site this results in a frequency vector of length *LJ*, and a covariance matrix of size *LJ × LJ*. By convention, reference sequence alleles have selection coefficients of zero, so the mutant allele selection coefficients at each site are normalized by subtracting the inferred coefficient for the reference allele.

### Branching process simulations

We implemented the superspreading branching process for the number of infected individuals in Python. We used a negative binomial distribution for the number of secondary infections caused by a group of individuals infected with the same pathogen variant. To test how finite sampling affects model estimates, we sampled *n*_*s*_ genomes per time point to use for analysis. We computed the single and double mutant frequencies, *x*_*i*_ and *x*_*ij*_, respectively, from the sampled sequences and estimated the selection coefficients from these using (1), possibly extended to account for multiple outbreaks or multiple alleles at each locus as described above. For the analysis of how finite sampling affects estimates, shown in **Supplementary Fig. 11**, we use the full version of the estimator given by (6). For all other simulations, we assume that the parameters *n, k*, and *R* are not known for inference and so we use the simplified estimator in (9) for inferring selection.

### Regions and time-series for SARS-CoV-2 analysis

We used sequence alignments and metadata downloaded from GISAID (ref. ^90^) on January 26, 2024, which includes more than 7.4 million sequences. One potential caution in interpreting this data is that not all sequences in the database will have been generated from unbiased surveillance efforts. Ideally, we would like to divide this data into the smallest separate areas that have outbreaks that are largely independent of those in the surrounding regions, so as to avoid biases due to travel between regions or unequal sampling in different locations. However, this needs to be balanced with the limitations of the data, since regions with poor sampling could contribute more noise than signal. We therefore divided data into the smallest regions available in the metadata that are still large enough such that infections resulting from travel outside of the region are likely to be far less frequent than transmission within the region. This results in the inclusion of mostly separate countries in Europe, states in North America, and a combination of countries and states in South America and Asia – dependent upon the size of the location. Two exceptions to this are that we separate northern and southern California due to the geographical separation of population centers, and we separate Northern Ireland from the rest of the United Kingdom due to its geographical isolation.

To minimize the effects of sampling noise, we chose regions and time-series within these regions based on the following criteria:

1. In any period of 5 days within the time-series there are at least 20 total samples.
2. The number of days in the time-series is greater than 20.
3. The number of new infections per day is at least 100.

The last criterion ensures that there are enough infected individuals that transmission is not driven overwhelmingly by stochasticity. We assessed the number of newly infected individuals by using the estimates provided by the Institute of Health Metrics and Evaluations ^91^. Since the dates provided in their estimates correspond to dates when individuals were infected, and dates in the GISAID sequence data correspond to dates when individuals were sequenced, we shifted the dates in the IHME data 5 days forward to roughly compensate for delays between infection and sequencing. We then eliminated days on which the estimated number of new infections was smaller than 100.

Our results are robust to reasonable variation in these parameters. Comparing the number of locations used and the sample sizes shown in **Supplementary Fig. 12** in the data to those used in the simulations shown in **Supplementary Fig. 1**, we expect our inference to accurately distinguish beneficial, deleterious, and neutral SNVs from one another.

### Data processing

We perform a number of preprocessing steps to ensure data quality. We first eliminated incomplete sequences with gaps or ambiguous nucleotides at more than 1% of the genome. We then removed sites from our analysis where gaps are observed at *>* 95% frequency, since these sites may represent very rare insertions or sequencing errors. We also removed sites in noncoding regions of the SARS-CoV-2 genome and ones where all observed SNVs are synonymous. We imputed gaps that are not associated with known variants and ambiguous nucleotides with the nucleotide at the same site that occurs most frequently in other sequences from the same region.

For the remaining sites, in each region we excluded rare SNVs whose frequency is not larger than 1% for at least 5 consecutive days. These sites, if included, are almost always inferred to have extremely small selection coefficients. Furthermore, since their frequencies are so small, their covariance with other sites is also small and is therefore unlikely to have a large effect on inference. We verified that different reasonable values for these cutoffs result in essentially identical selection coefficients (**Supplementary Fig. 13**).

### Calculating frequency changes and covariances

To increase robustness to finite sampling in time, we integrated terms in (6) and other time-dependent equations over time by assuming that frequencies are piecewise linear, rather than summing contributions from each time point ^76^. This results in diagonal terms of the integrated covariance given by

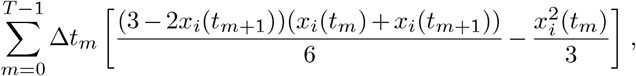

and off-diagonal elements

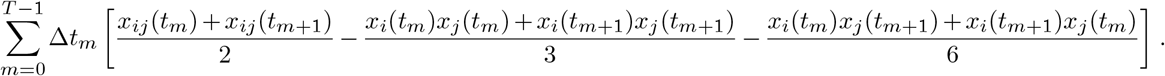

For obtaining reliable estimates of the changes in SNV frequencies (the term *x*(*t*_*T*_)− *x*(*t*_0_) in (8)), it is important to have enough sequences to avoid large errors due to finite sampling. On the other hand, if a large number of days are used at the end or the start of the time-series to calculate the frequencies, then the frequency changes are likely underestimates. To balance these competing issues, we calculated ***x***(*t*_*T*_) as the frequencies in the window of the final 15 days and ***x***(*t*_0_) as the frequencies in the window of the first 15 days for each time-series and region with poor sampling. This smoothing is necessary especially in regions where sampling is sparse, where the number of genomes sampled on a particular day may be as small as 1 or 2. If there are at least 200 sampled sequences in a period of less than 15 days at the start or the end of the time-series, then the window size was taken as the smallest number of days in which there was a total of at least 200 sequences. We confirmed that our results are robust to reasonable changes of this window size of 15 days (**Supplementary Fig. 13**).

We also normalized time in units of serial intervals or “generations” by dividing the integrated covariance matrix by 5, following results that the serial interval for SARS-CoV-2 is roughly 5 days ^92–94^. This allows us to convert from units of time in days to generations, as in (8).

### Calculating selection coefficients

After the above preprocessing there remain 1,320 SNVs observed at a frequency above 1% for at least 5 consecutive days in at least one region and observed at least 5 times. We assume constant values for *R, n*, and *k* in all regions, and use (9) to estimate selection. When *R, n*, and *k* are constant, these terms can be effectively absorbed into the regularization *γ′*.

We normalize selection coefficients such that the nucleotide for the WIV04reference sequence at each site has a selection coefficient of 0. To do this, we subtract the selection coefficient for the reference nucleotide from the inferred coefficient for each other allele at that site after all selection coefficients have been computed.

We used these estimates for the selection coefficients for nonsynonymous SNVs to estimate the corresponding selection coefficients for amino acid substitutions (**Table 1**). If there were multiple SNVs in a codon that result in the same amino acid variant, but are not strongly linked to one another, then the selection coefficient for the amino acid was calculated as the largest (in absolute value) of the SNVs. If there were multiple SNVs in the same codon that yield the same amino acid and these SNVs are strongly linked to one another, then the selection coefficient for the mutant amino acid was calculated as the sum of the selection coefficients for the SNVs. Our reasoning behind this choice is that selection coefficients that are extremely close to zero are mostly for alternative nucleotides that are observed very infrequently in the data, and so the inferred coefficients for these nucleotides are unlikely to reflect the typical effects of a given mutation.

We calculated selection coefficients for major variants by summing the individual nucleotide SNVs that define the variant, which follows from our assumption of additive fitness. The SNVs for major named variants such as Alpha and Delta were identified according to the mutations provided by https://covariants.org. Results of this analysis are shown in **Figs. 2-3, Supplementary Figs. 3-5** and **14-15**, and **Supplementary Table 1. Supplementary Figs. 14-15** quantify uncertainty in the inferred selection coefficients, based on both theoretical uncertainty in the selection coefficient estimator and finite sampling noise. For a detailed discussion, see Supplementary Information.

### Computational complexity

Here we briefly discuss the computational complexity of our method. The steps in our data processing are:

1. Clean the data (eliminate sequences with large numbers of Ns or gaps, etc.).
2. Separate the data by time and region.
3. Identify SNVs observed above the minimum frequency threshold.
4. Compute SNV covariance matrices/changes in SNV frequencies in each region and integrate them over time.
5. Infer the selection coefficients, which involves inverting the total integrated SNV covariance matrix.

Let *L* be the length of the SARS-CoV-2 sequence (roughly 3 *×* 10^4^ bps) and let *M* be the total number of sequences (roughly 10^7^ including data taken up until January 26th, 2024). Then, steps 1 and 2 involve computations that scale as 𝒪 (*M*). Step 3 is 𝒪 (*ML*). This step also introduces a new parameter relevant for the scaling of the problem, which is the fraction of SNVs that are observed at high enough frequencies to be included in our analysis. Let us call this fraction *p*, which is roughly 0.35 with our current settings. Naively, step 4 then involves a computation that scales like 𝒪 (*M* (*pL*)^2^). However, the calculation of the covariance can easily be parallelized across regions. In each individual region, the fraction of SNVs that are observed at high enough frequencies to be included is a different parameter *q* and the number of sequences in the region is a parameter *M*_*r*_. The largest *q* that we find in the regions analyzed is around 0.05. For *N*_*r*_ separate regions (149 in our analysis), step 4 then involves *N*_*r*_ parallel computations that scale like 𝒪 (*M*_*r*_(*qL*)^2^). Due to the matrix inversion, step 5 requires 𝒪 ((*pL*)^3^) computations to complete.

### Choice of regularization

In principle, the regularization strength *γ′* is related to the width of the prior distribution for SNV selection coefficients. The regularization strength also plays a role in reducing noise in selection coefficient estimates due to finite sampling of viral sequences. This is especially important for SNVs that are observed only briefly in data, as they will have small integrated variances in the “denominator” of (6). Larger values of the regularization more strongly suppress noise, but they also shrink inferred selection coefficients towards zero.

We use a regularization strength of *γ′* = 40. For much smaller values of *γ′*, selection coefficient estimates are unstable due to sampling noise. However, inferred selection coefficients stabilize and become insensitive to the precise value of *γ′* for *γ′* ≥10 (**Supplementary Fig. 13**). Larger values of *γ′* will result in selection coefficients with smaller absolute values, but for large enough *γ′* the rank ordering of inferred selection coefficients is highly reliable. In summary, the coefficients that appear to be the most beneficial or deleterious remain this way regardless of reasonable choices for *γ′*, though their precise values scales with the regularization strength.

### Identification of HG SNVs

To estimate how quickly we can detect a transmission advantage for a new SNV or variant, and to explore the sensitivity of this detection, we inferred selection coefficients for all variants *ŵ*(including SNVs and collections of SNVs that are strongly linked to one another), for every day in every region separately. To determine sets of strongly linked SNVs, we considered the following statistics. If the number of genomes with a SNV at site *i* is called *h*_*i*_ and the number of genomes with SNVs at both site *i* and site *j* is *h*_*ij*_, then we say that two sites *i* and *j* are strongly linked if *h*_*ij*_*/h*_*i*_ and *h*_*ij*_*/h*_*j*_ are both greater than 80%.

To form sets of strongly linked SNVs, we combined all pairs of strongly linked SNVs that share SNVs in common. For example, if SNV *i* is strongly linked with SNV *j*, and SNV *j* is strongly linked with SNV *k*, then {*i, j, k*} forms one set of strongly linked SNVs. With the frequency cutoff that we have used for the definition of strongly linked SNVs (80%), the great majority of SNVs in each set of strongly linked SNVs are strongly linked to all other SNVs in the same set. We computed selection coefficients for sets of strongly linked SNVs by summing the contributions from individual SNVs.

Data was trimmed by submission date such that the selection coefficients for a specific day are calculated using only sequences that were submitted to GISAID on or before that day. We then progressively step through time in each region, adding newly submitted sequences and reanalyzing the data again. At each time point in every region, groups of strongly linked SNVs are recalculated using the method described above and selection coefficients for the collections are computed again. To compare the HG SNVs with well-studied major SARS-CoV-2 variants, which are widely understood to have a significant transmission advantage relative to ancestral SARS-CoV-2, we performed this analysis using data from the beginning of the pandemic through June 2022.

As described in the main text, we suspect that collections of SNVs with large inferred selection coefficients are much more likely to exhibit real advantages in transmission. Therefore, we used a classification scheme where variants with selection coefficients *ŵ* > *θ* for some cutoff *θ* are classified as “high growth (HG)” variants. At each time step, we removed any SNVs that were classified as HG from all future analyses in that region. In this way, any SNV can only contribute to the detection of a single variant in a region (e.g., for a mutation that belongs to both Alpha and Omicron, if the mutation was labeled as HG during the rise of Alpha in a given region, then that mutation will not be considered when analyzing later Omicron sequences in the same region).

After a mutation is detected in a region, we also remove all other nucleotide mutations at that site from future analysis in the region. The reason for this is the following. The choice of a normal prior distribution on the selection coefficients enforces that the sum of the selection coefficients for a specific site is zero. We then re-normalize the selection coefficients so that the selection coefficient for the WIV04 reference nucleotide is set to zero. This is done by subtracting its value from the selection coefficients for all other nucleotides at that site, as described above. In the ordinary situation where only two different nucleotides are observed at a site, this normalization procedure results in the apparent inflation of selection coefficients for unobserved nucleotides at the same site. If one of these other nucleotides is later observed at a low frequency, this could result in an incorrect detection. For this reason, we remove all nucleotides at the same site from consideration in a region after any single nucleotide has been detected.

We performed inference for the detection of HG variants across each region individually, as the same new variant is unlikely to first appear at identical times in multiple regions. This limits the strength of statistical information to infer selection because information is not aggregated across regions. For this reason, we used a lower regularization of *γ* = 10 for regional analysis to prevent the strong suppression of inferred selection coefficients. Tuning the threshold of detection *θ* allows one to adjust the tradeoff between noise, which may lead to false positives, and detection speed. Results of this analysis are presented in **Supplementary Figs. 6-7**. The analysis shown in **Fig. 4** uses an analogous approach where selection coefficients were computed over time for Alpha, Delta, and Omicron (BA.1) SNVs in specific regions, but without the additional step of classifying SNVs as HG.

To succinctly visualize HG SNVs linked with major variants (**Supplementary Fig. 7**), we grouped the regions into 7 broad categories, allowing for clearer trend analysis. For each major variant within these broad regions, we identified HG groups with associated mutations and plotted the cumulative fraction of variant-defining mutations over time. Data regarding variant-defining mutations was sourced from https://covariants.org.

### Data and code

Sets of processed data, computer code, and scripts that we have used in our analysis are available in the GitHub repository located at https://github.com/bartonlab/paper-SARS-CoV-2-inference. This repository also contains Jupyter notebooks that can be run to reproduce the results presented here, using sequence data and metadata from GISAID. A full list of originating and submitting laboratories for the sequences used in our analysis can be found at https://www.gisaid.org using the EPI-SET-ID: EPI_SET_240815xt.

---

**Supplementary Fig. 1.**
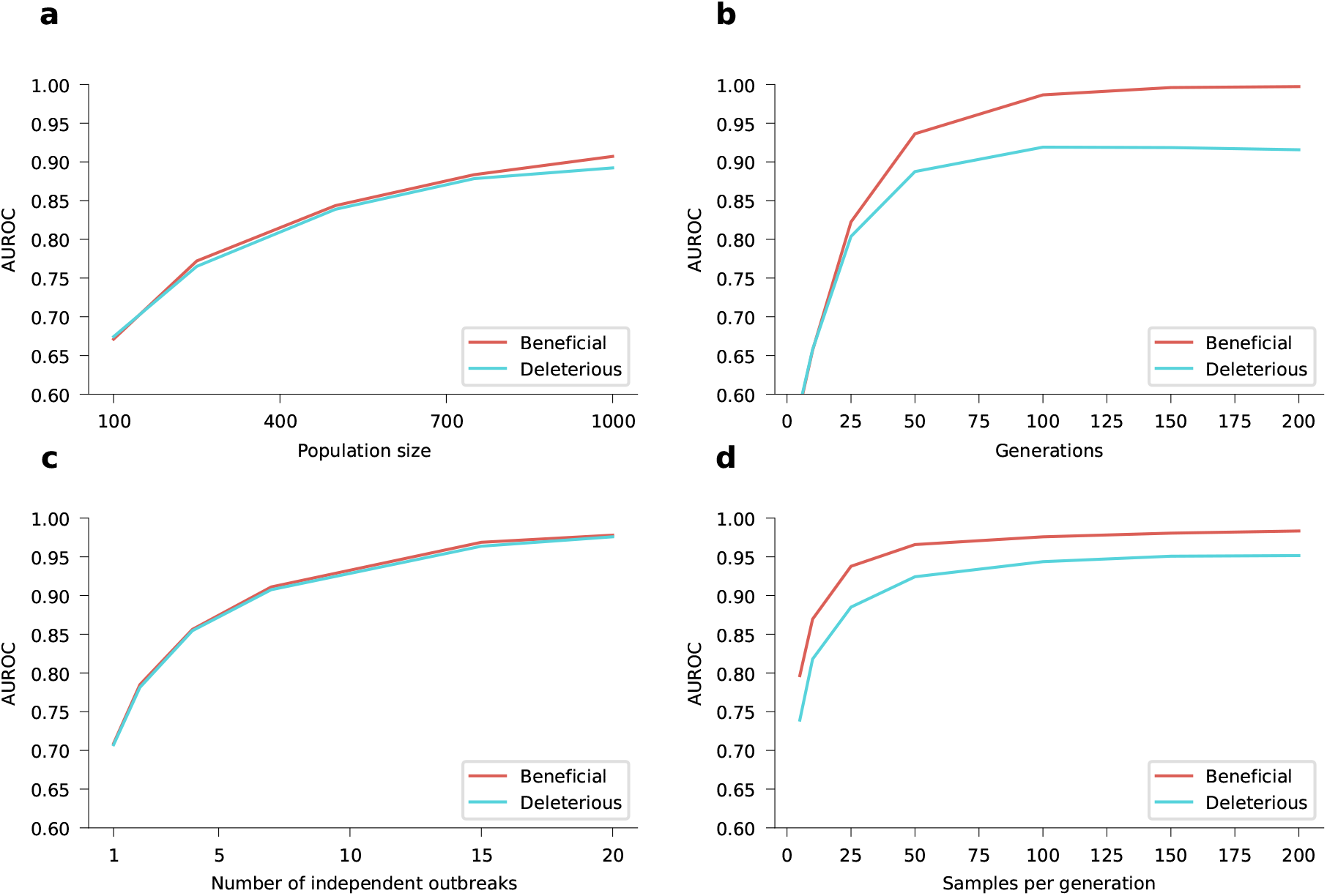
Accuracy of inference for different parameters. How the AUROC scores for both beneficial SNVs (in red) and deleterious SNVs (in blue) depends upon the different model parameters. **a**, Inference accuracy for different values of newly-infected population size. The parameters used are 10 simulations each with 50 sampled genomes per generation for 25 generations. **b**, Inference accuracy for different numbers of generations (serial intervals). Data is from a single simulation with 25 samples per generation and a newly-infected population size of 10,000. **c**, Inference accuracy for different numbers of independent outbreaks (simulations). The parameters used are 50 samples per generation for 10 generations and a newly-infected population size of 10,000. **d**, Inference accuracy for different values of samples per generations. Data is from a single simulation with 50 generations with a newly-infected population size of 10,000. The initial population is a mixture of two variants with beneficial SNVs (*s* = 0.03), two with neutral SNVs (*s* = 0), and two with deleterious SNVs (*s* = −0.03). Dispersion parameter *k* is fixed at 0.1. This is the same initial population composition as described in **Fig. 1**. All AUROC scores are calculated by averaging over 1,000 replicate simulations.

**Supplementary Fig. 2.**
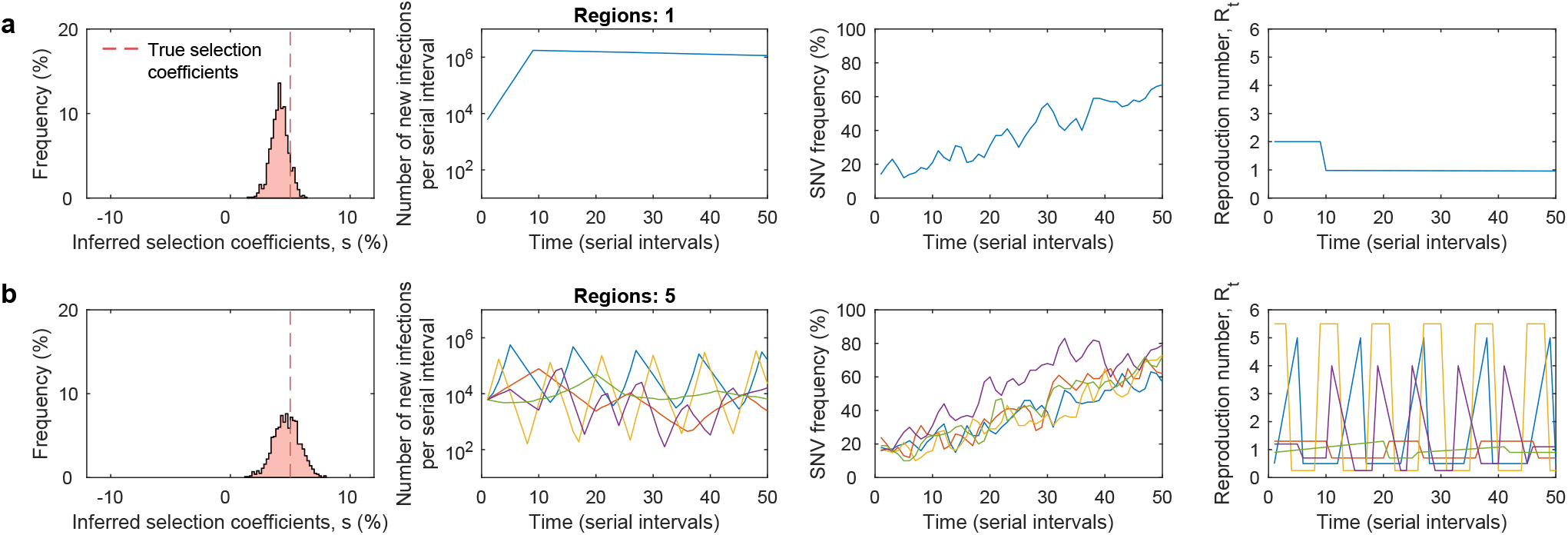
Inference is robust to variation of reproduction number, *R*, across regions. Our approach provides a systematic way to combine data from outbreaks in multiple regions. Simulations show that the estimator in (9) has good performance whether the selection coefficients are inferred based on data from, **a**, a single region or, **b**, five regions. *Simulation parameters*. The initial population in each region is a mixture of a neutral variant with no mutations and a variant with a beneficial SNV (*s* = 0.05). The same beneficial SNV appears in all 5 regions. Each region has a different profile of the time-varying reproduction number, *R* (rightmost panel). In the first simulation, the number of newly infected individuals per serial interval rises rapidly from 6,000 to around 10,000 and stays nearly constant thereafter. While in the second simulation it has a different profile for each region, all the while staying between 100 and 100,000. Dispersion parameter *k* is fixed at 0.1 for both simulation scenarios.

**Supplementary Fig. 3.**
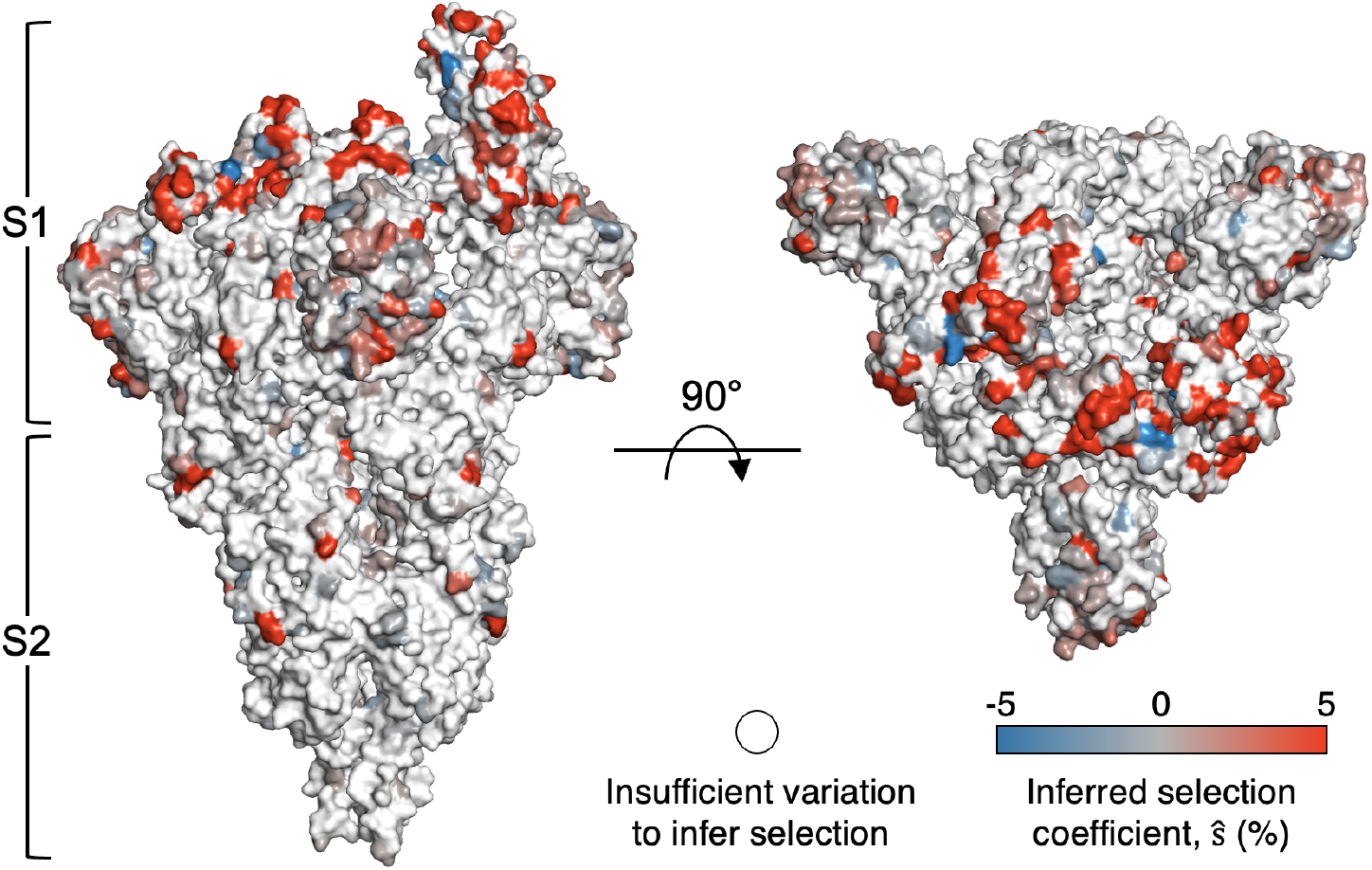
Inferred selection coefficients for Spike mutations mapped on the crystal structure. The majority of the inferred strongly selected mutations are in the S1 subunit of Spike. For sites with multiple mutations, the mutation with the largest magnitude of inferred selection coefficient was used for mapping. Structure of the Spike protein was obtained from http://rcsb.org/ (PDB ID: 7WG7) (ref. ^95^).

**Supplementary Fig. 4.**
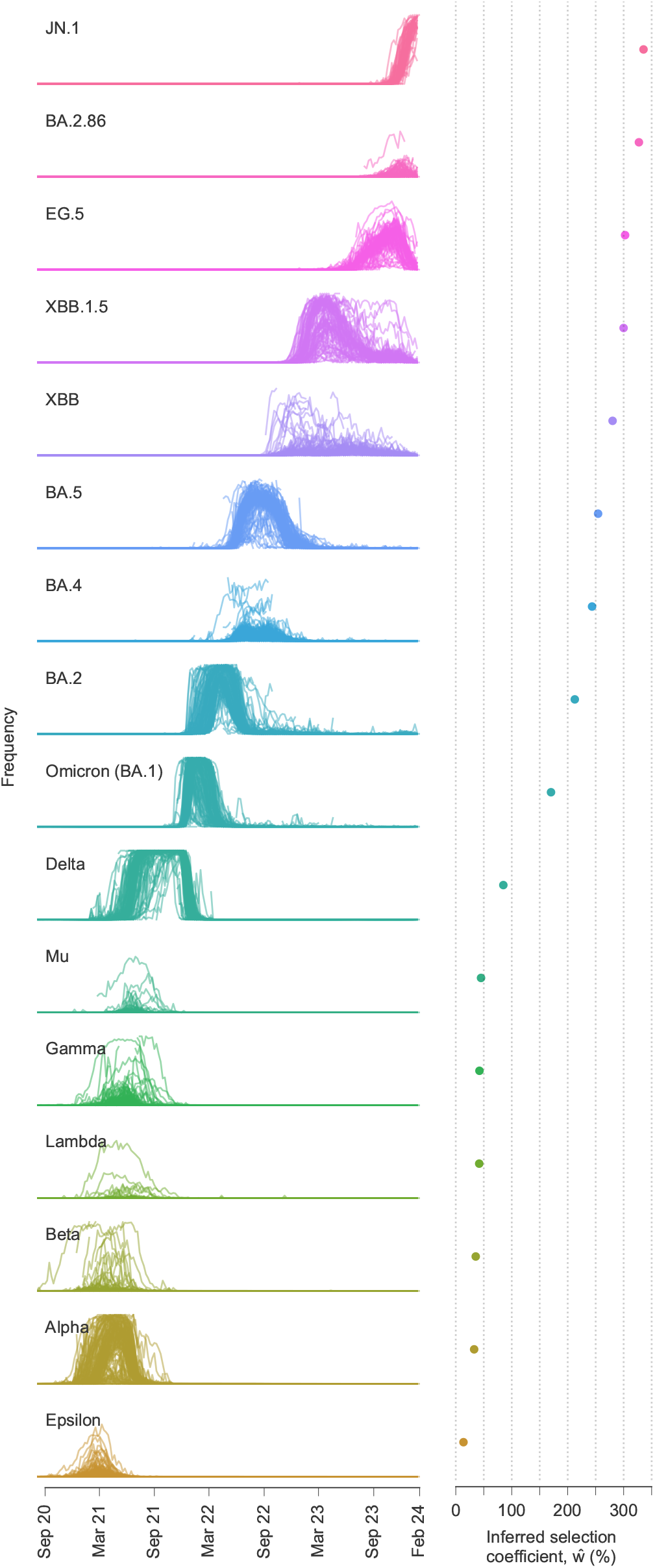
Multiple SARS-CoV-2 variants strongly increase transmission rate. Frequencies of major variants and their total inferred selection coefficients, shown as mean values *±* one s.d. from bootstrap subsampling of regional data (Methods), defined relative to the WIV04 reference sequence. Selection coefficients for variants with multiple SNVs are obtained by summing the effects of all variant-defining SNVs. Because our method uses global data and accounts for competition between variants, we infer large transmission advantages even for variants such as Gamma, Beta, Lambda, and Epsilon, which never achieved the same level of global dominance as variants such as Alpha and Delta.

**Supplementary Fig. 5.**
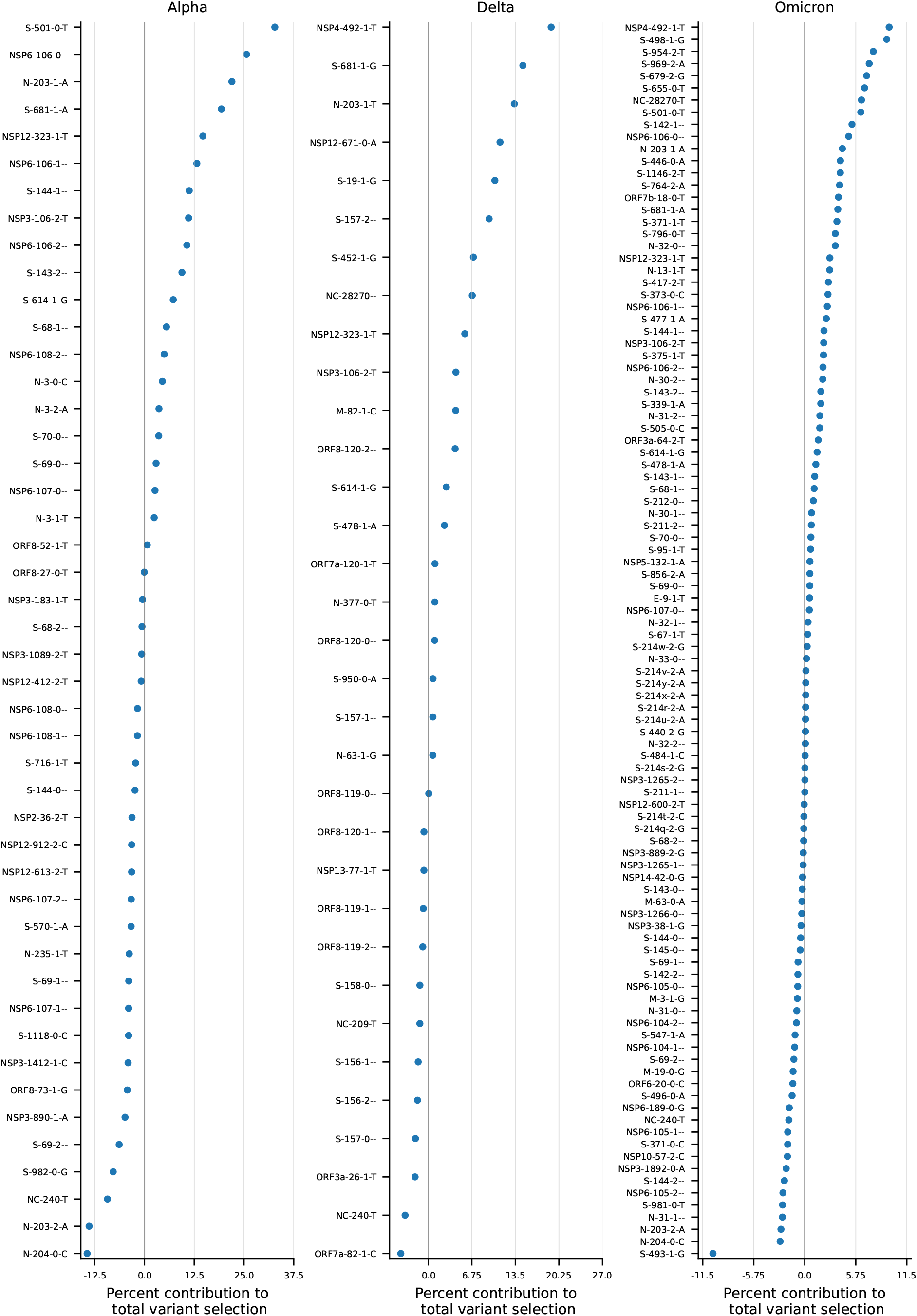
For major variants, a minority of SNVs provide most of the total increase in transmission. Fraction of the total increase in transmission for Alpha, Delta, and Omicron (BA.1) provided by each variant-defining mutation. For each variant, a few strongly beneficial mutations provide most of the total increase in transmission. Most other mutations are inferred to be nearly neutral. For some variants, a small number of mutations are inferred to be substantially deleterious.

**Supplementary Fig. 6.**
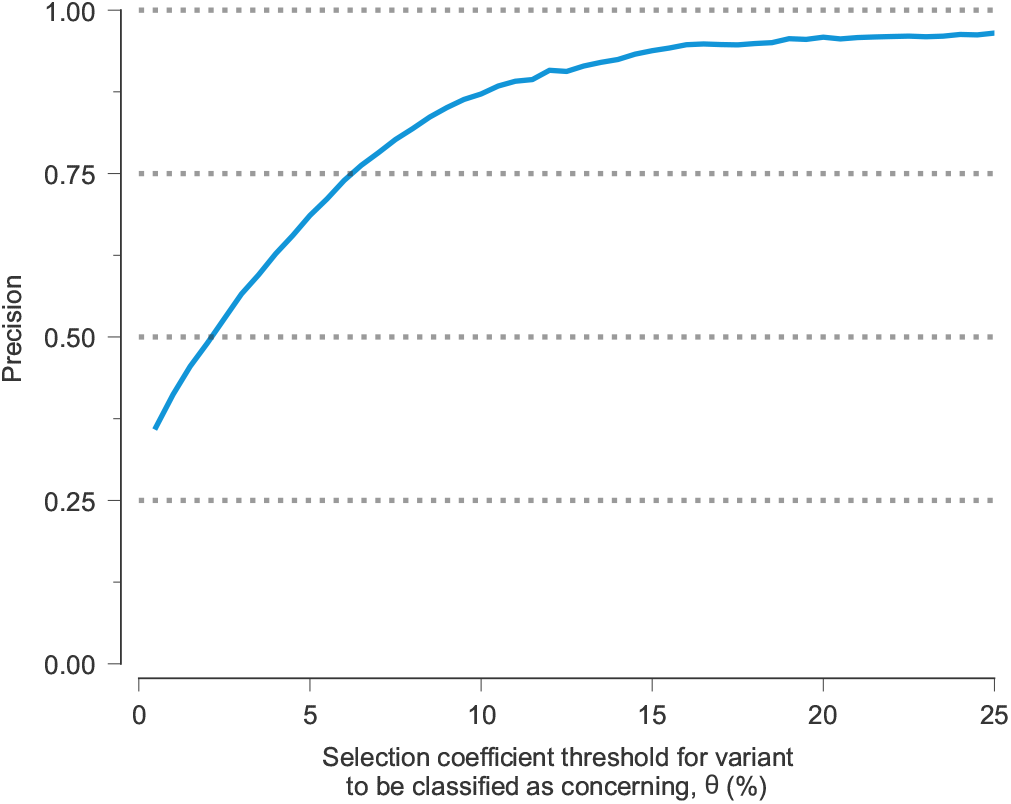
Variants with large inferred selection coefficients are overwhelmingly likely to belong to major variants, even when selection is estimated as data becomes available. Fraction of variants classified as concerning with SNVs that belong to major SARS-CoV-2 variants, plotted as a function of the selection coefficient threshold *θ* used for classification. We consider (groups of) SNVs classified as concerning to be true positives if they belong to major variants and false positives otherwise. With this definition, this fraction is equivalent to the precision for classification.

**Supplementary Fig. 7.**
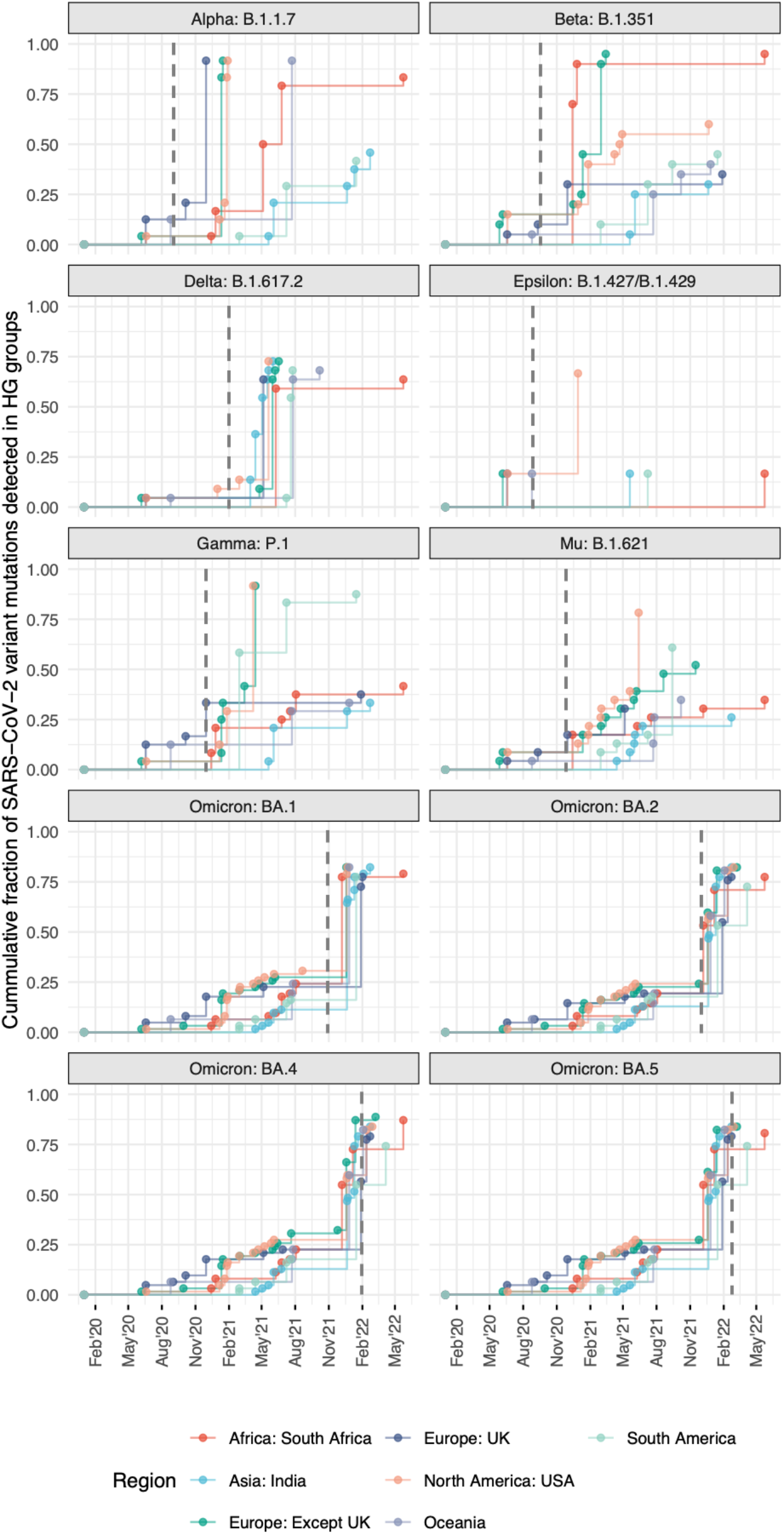
Cumulative fraction of SARS-CoV-2 variant-defining mutations identified as HG across regions. Results are shown for 10 major variants across 7 broad geographical regions. The vertical dashed line indicates the earliest sample date for each variant. Data of variant-defining mutations and their earliest sample dates were obtained from https://covariants.org.

**Supplementary Fig. 8.**
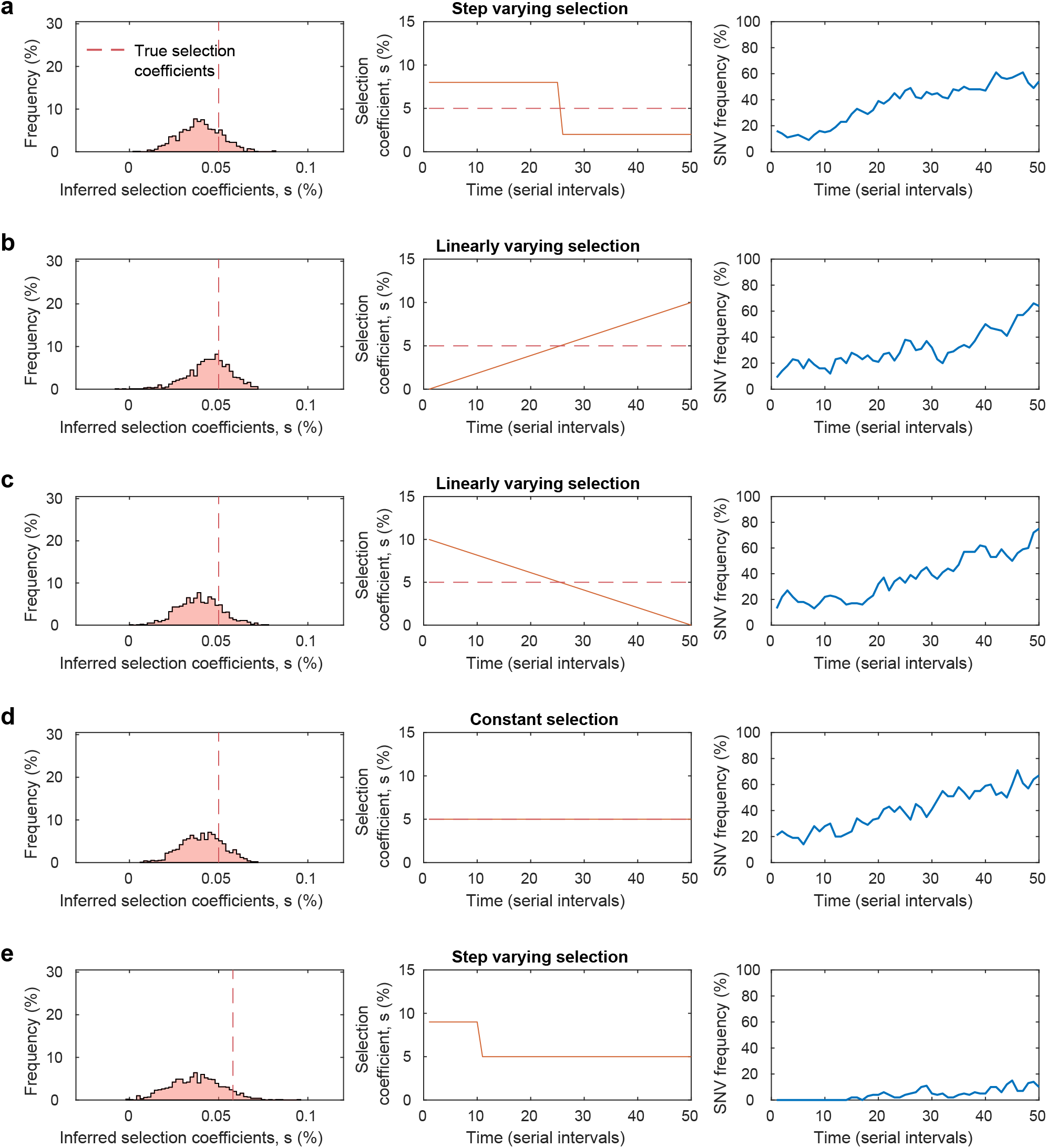
Average value inferred for time-varying selection coefficients. We simulated five scenarios of time-varying selection coefficients: **a**, step varying, **b**, linearly increasing, **c**, linearly decreasing, **d**, constant over time and, **e**, step varying where the SNV appears in the population after the true selection coefficient has changed. In each case, the inferred selection coefficient is close to the average of the time-varying selection coefficient over the time when the SNV was present in the population. *Simulation parameters*. The initial population in the first four simulation scenarios is a mixture of a neutral variant with no mutations and a variant with a beneficial SNV with a time-varying selection coefficient (center panels). In the fifth simulation scenario, the initial population consists entirely of the neutral variant with the beneficial mutant appearing after 15 serial intervals. The number of newly infected individuals per serial interval rises rapidly from 6,000 to around 10,000 and stays nearly constant thereafter. Dispersion parameter k is fixed at 0.1 for all simulation scenarios.

**Supplementary Fig. 9.**
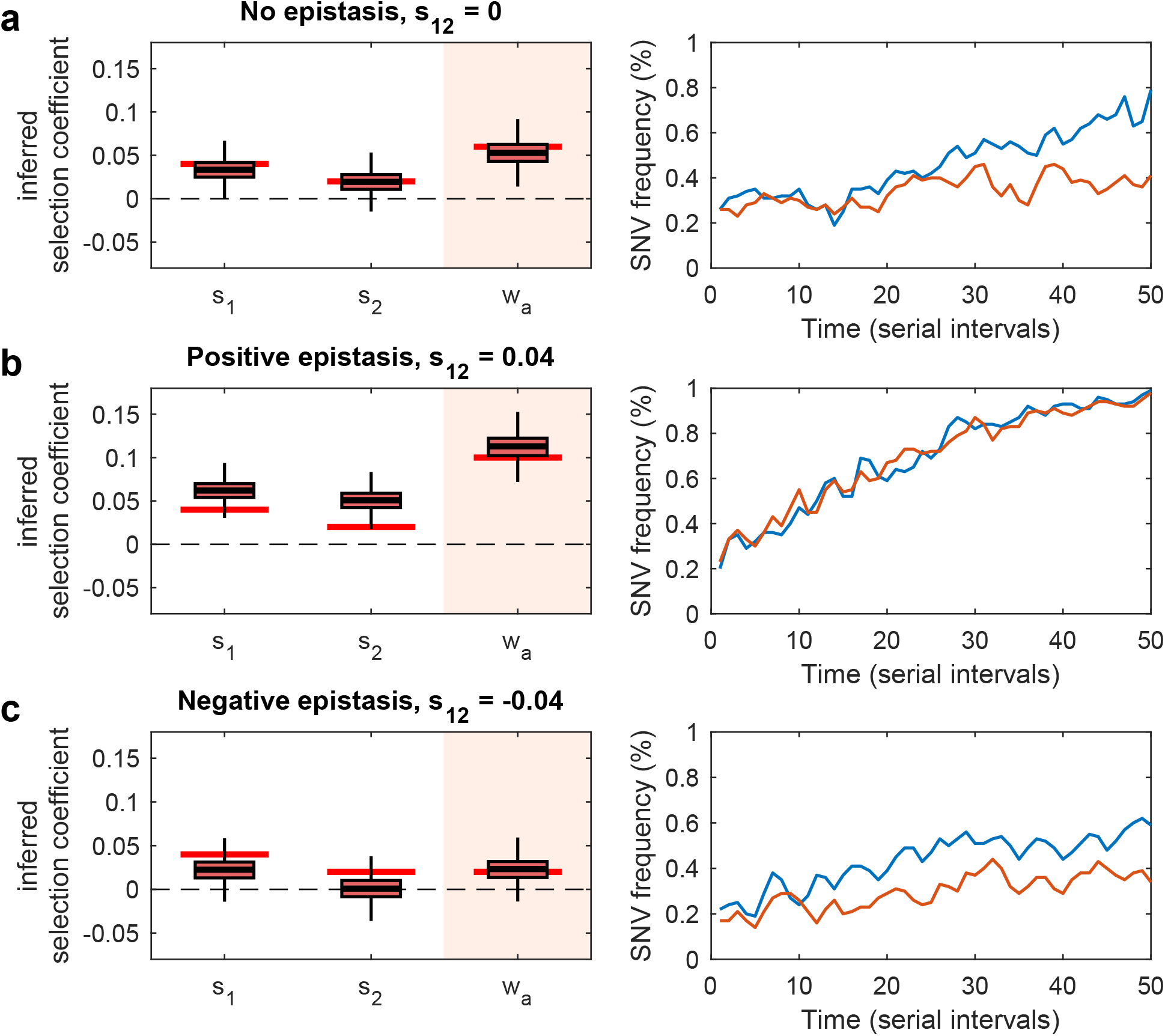
Accurate inference of variant fitness in the presence of epistasis. **a**, Both SNV selection coefficients and variant selection coefficients are inferred accurately in the absence of epistasis. Inferred selection coefficients over 1,000 runs are shown in box plots, with true values for the parameters shown with solid bars in red. The lower and upper edge of the box plot correspond to the 25th to 75th percentiles, the bar inside the box plot corresponds to the median while the top and bottom whiskers show the maximum and minimum value within 1.5 times the interquartile range. In scenarios with positive epistasis (**b**) or negative epistasis (**c**), our method attributes the effect of epistasis to selection coefficients. Thus, while the inferred SNV selection coefficients may be under- or over-estimated, the inferred variant selection coefficients are recovered. *Simulation parameters*. We simulate a two-locus system where the initial population consists of a mixture of all four variants, i.e., a neutral variant with no mutations, a variant with two beneficial SNVs (*s*_1_ = 0.04, *s*_2_ = 0.02), and both single SNV variants. The initial frequencies in the population of the neutral, the two single mutant variants, and the double mutant variants are set to 67%, 10%, 10%, and 13%. We simulate three scenarios with the epistasis term taking on values *s*_12_ = {0, 0.04, -0.04}. Here the selection coefficient for the double mutant is *s*_1_ + *s*_2_ + *s*_12_. The number of newly infected individuals per serial interval rises rapidly from 6,000 to around 10,000 and stays nearly constant thereafter. Dispersion parameter *k* is fixed at 0.1 for all simulation scenarios.

**Supplementary Fig. 10.**
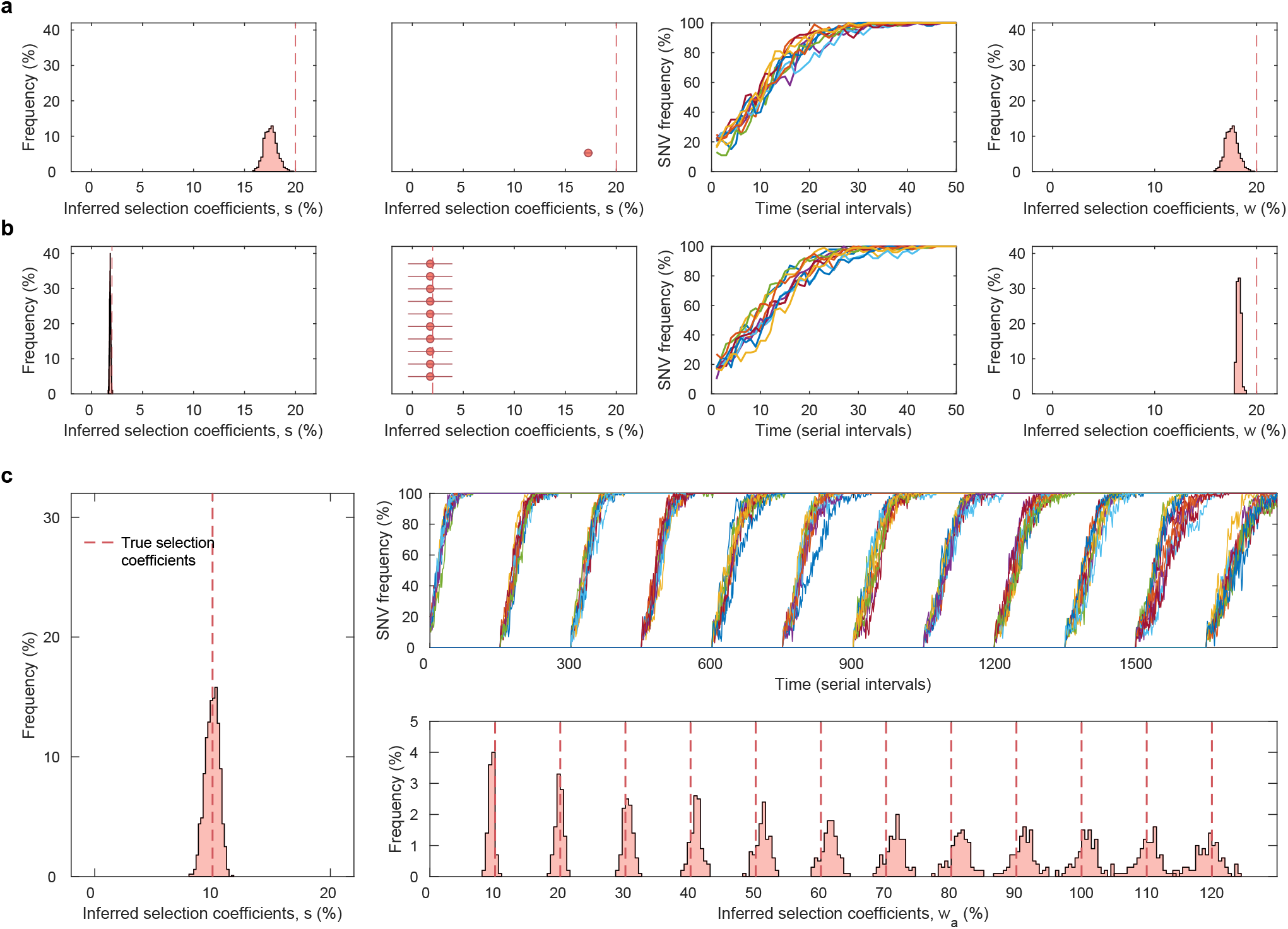
Ability to estimate large variant selection coefficients, *w*_*a*_. While the estimate (9) is derived assuming selection coefficients are small, simulations show that combining data from multiple regions allows for accurate estimation of both large SNV selection coefficients, *s*, and variant selection coefficients, *w*_*a*_. **a**, A scenario with a variant containing a single strongly beneficial SNV (*s* = 0.2) and, **b**, a scenario with a variant containing 10 mildly beneficial SNVs (*s* = 0.02). The true variant selection coefficient *w*_*a*_ has the same magnitude in both simulation scenarios (*w*_*a*_ = 0.2). **c**, Simulating a scenario where 12 beneficial SNVs (*s* = 0.1) appear and fixate successively (top right panel), such that *w*_*a*_ ranges from 0.1 to 1.2, both the SNV (left panel) and variant selection coefficient (bottom right panel) were estimated accurately. Results are obtained by combining data from 10 regions. Histograms are obtained from 1, 000 replicate simulations. *Simulation parameters*. In the simulation scenarios considered in **a** and **b**, the initial population in each region consists of a mixture of a neutral variant with no mutations along with a variant with a single strongly beneficial SNV (*s* = 0.2), or a variant with 10 beneficial SNVs (*s* = 0.02) respectively. In the simulation in **c**, each region’s initial population consists of a mixture of a neutral variant with no mutations along with a variant with beneficial mutations. In this latter variant, 12 beneficial mutations (*s* = 0.1) appear and fixate in succession such that the variant selection coefficient varies from *w*_*a*_ = 0.1 to *w*_*a*_ = 1.2. The same variant appears in 10 independent regions in all simulation scenarios. The number of newly infected individuals per serial interval is nearly constant around 10,000. Dispersion parameter *k* is fixed at 0.1.

**Supplementary Fig. 11.**
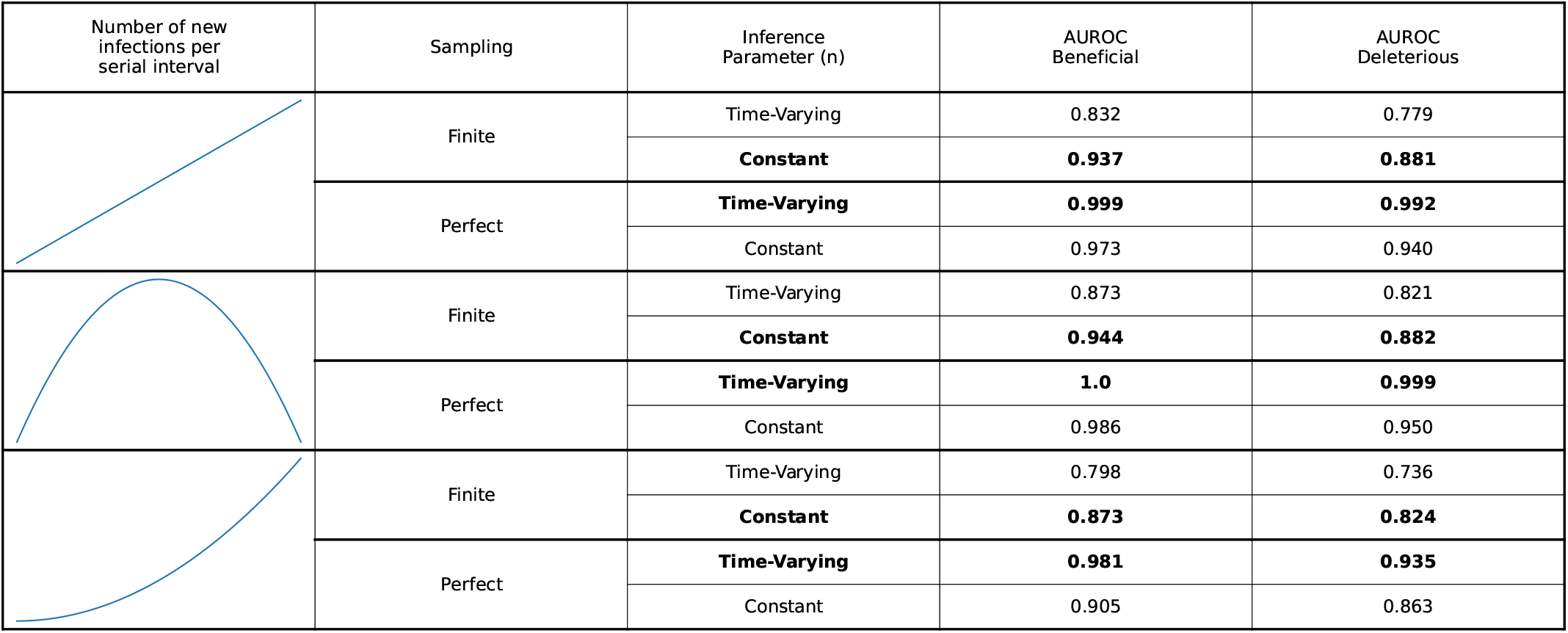
Effects of finite sampling on inference using constant and time-varying parameters. The ability of the model to distinguish beneficial and deleterious SNVs, as measured by the AUROC score, depending on whether the sampling is perfect or finite and whether constant parameters or the true time-varying parameters are used for the number of new infections per serial interval *n* in the inference. If parameters are considered to be constant, then these parameters are not required for inference using (8). Both simulations use constant values of *k* = 0.01 and *R* = 1. The results are similar but less dramatic if the correct time-varying values are used for *k* or *R* as well. Results are shown for different trajectories of numbers of infections and are consistent regardless of the trajectory. In the upper panel, the number of new infections per serial interval, *n*, starts at 5, 000 and rises linearly to 100, 000. In the middle panel, *n* starts at 10, 000, rises quadratically to a maximum of 200, 000, and then falls back to the original number. In the final panel, *n* rises from an initial size of 1, 000 to a final size of 65, 000. All simulations are run for 50 serial intervals. Rows that yield better inference are marked by bold text. If sampling is finite, then it is better to use constant parameters; if sampling is perfect, then it is better to use the real time-varying parameters. The initial population of individuals are infected with a mixture of two variants with beneficial SNVs (*s* = 0.03), two with neutral SNVs (*s* = 0), and two with deleterious SNVs (*s* = −0.03), as in **Fig. 1**. Simulations are run for 50 simulations with 25 samples in each serial interval, and AUROC scores are averaged over 1,000 replicate simulations.

**Supplementary Fig. 12.**
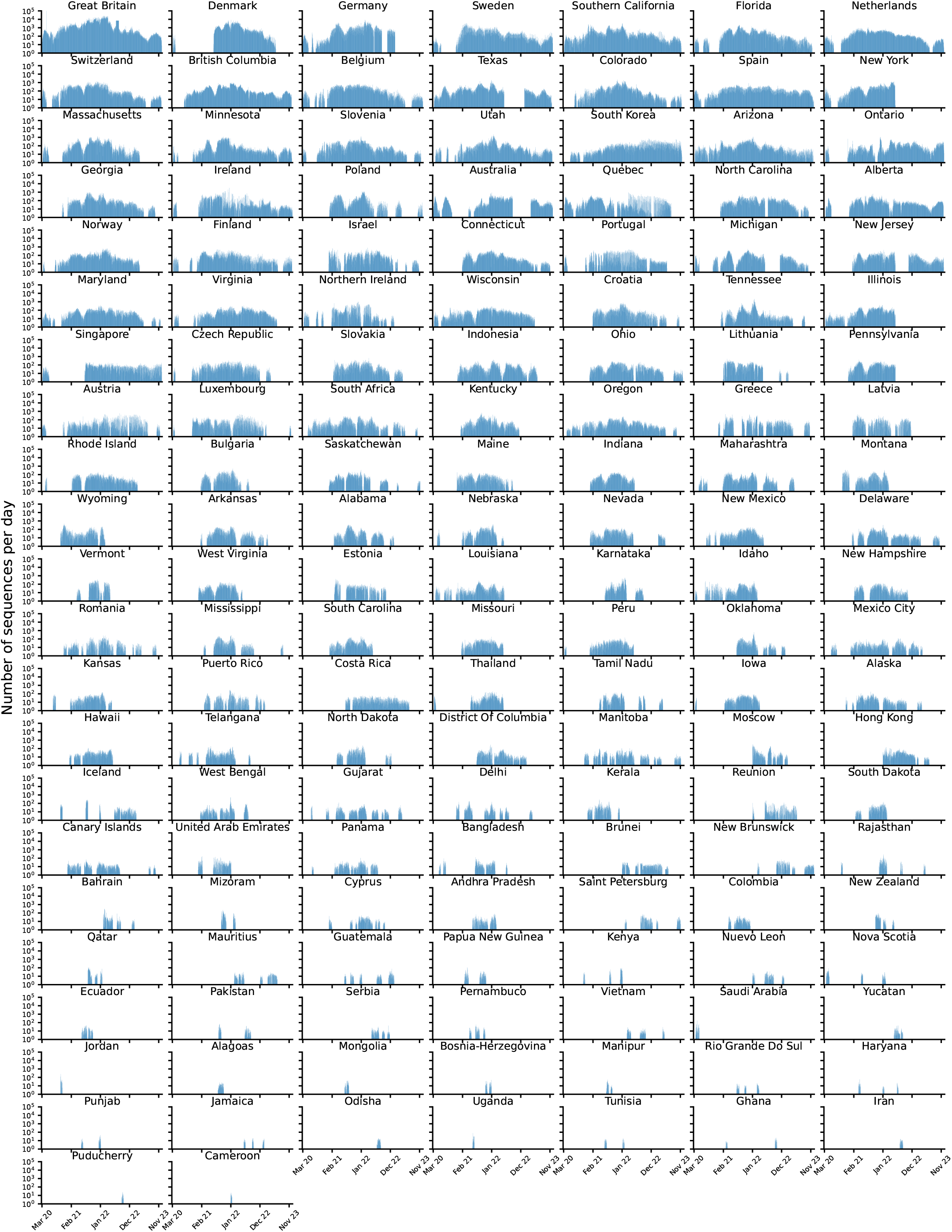
Sampling Distributions. The number of genomes per day in the regions that are used for inference.

**Supplementary Fig. 13.**
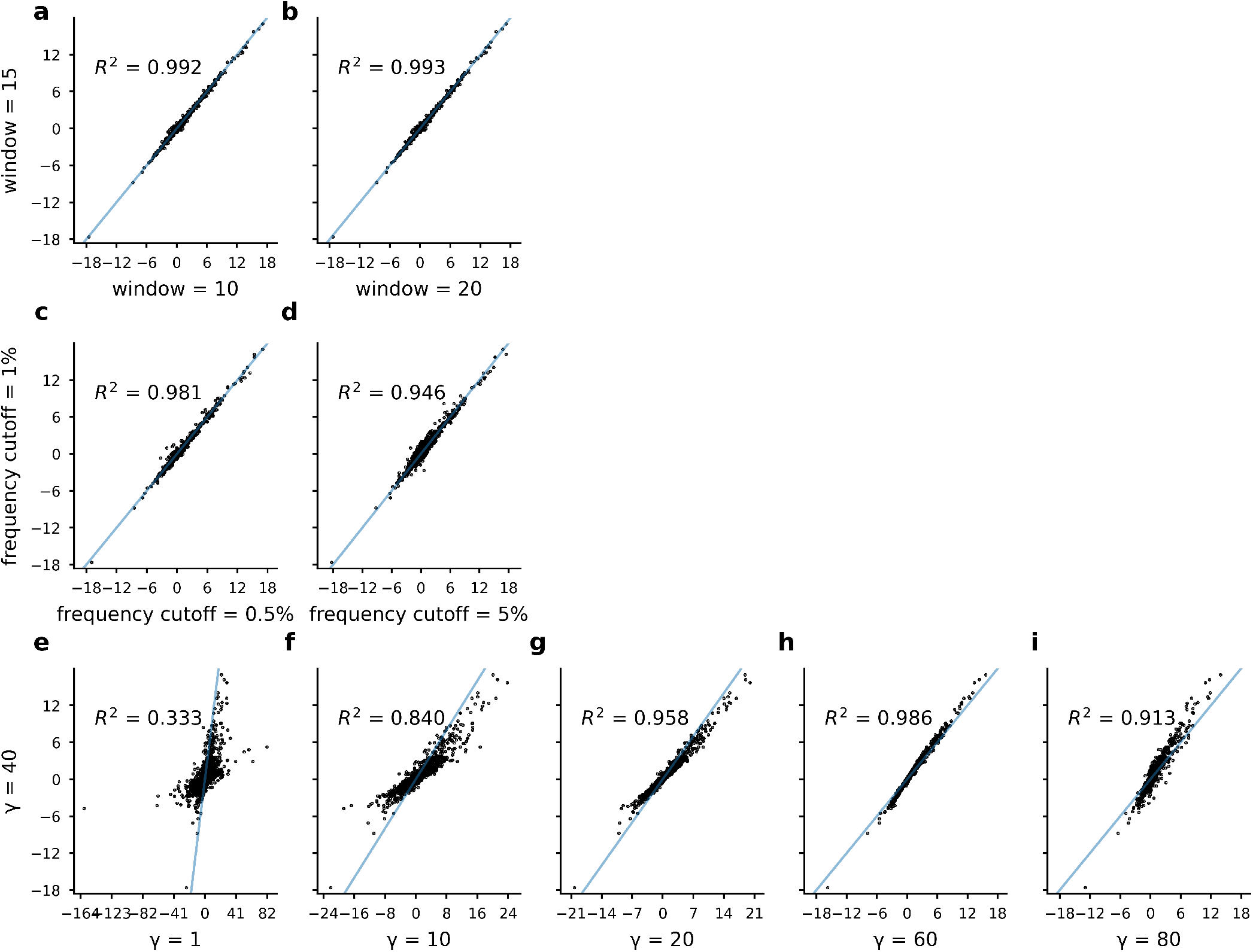
Inferred selection coefficients are robust to different values of the regularization *γ*′, different frequency cutoffs, and different numbers of days used to calculate the frequency changes. **a-b**, Comparison of inferred coefficients when the number of days at the beginning and end of the time-series are used in order to calculate the frequency changes. Inferred coefficients are largely robust to these changes **c-d**, Comparison of inferred coefficients for different frequency cutoffs. Including more or less sites does not alter the order of inferred coefficients. **e-i**, Comparison of inferred coefficients for different values of the regularization. Altering the regularization value has little effect upon the distribution of inferred selection coefficients, and selection coefficients for different values of the regularization are highly correlated.

**Supplementary Fig. 14.**
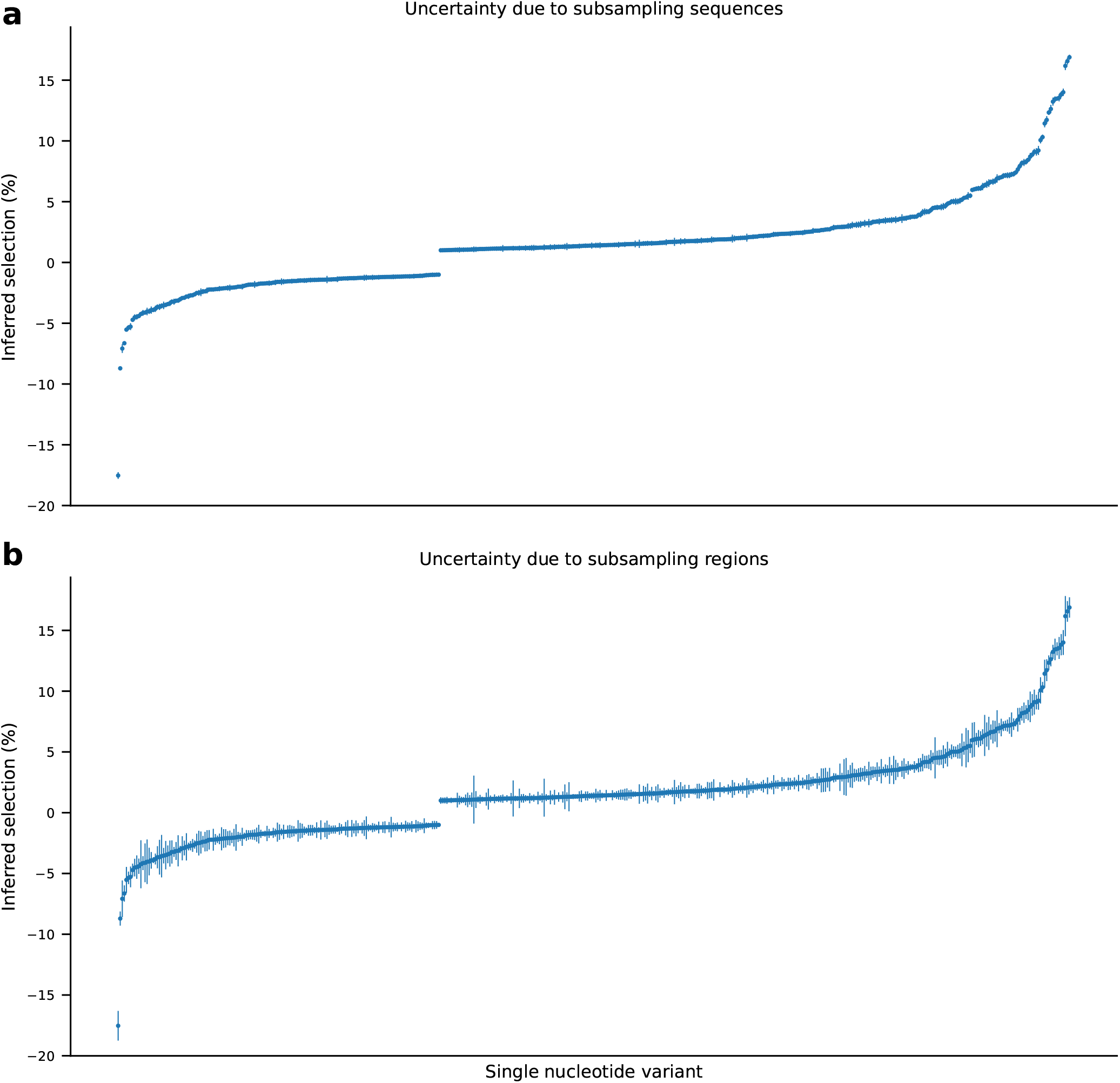
Selection coefficient estimates and uncertainty. Plots of all inferred selection coefficients with absolute values greater than 1%. **a**, Selection coefficients with uncertainty estimates from bootstrapping the sequences in each region. 20 sequences were sampled per time point per region, with replacement. Error bars represent standard deviations of the inferred coefficients computed over 100 bootstrap samples. **b**, Selection coefficients with uncertainty estimates from subsampling the regions used. For each run, we inferred selection coefficients using a random subsample of 80% of the total number of regions. Error bars represent standard deviations of the inferred coefficients computed over 100 samples.

**Supplementary Fig. 15.**
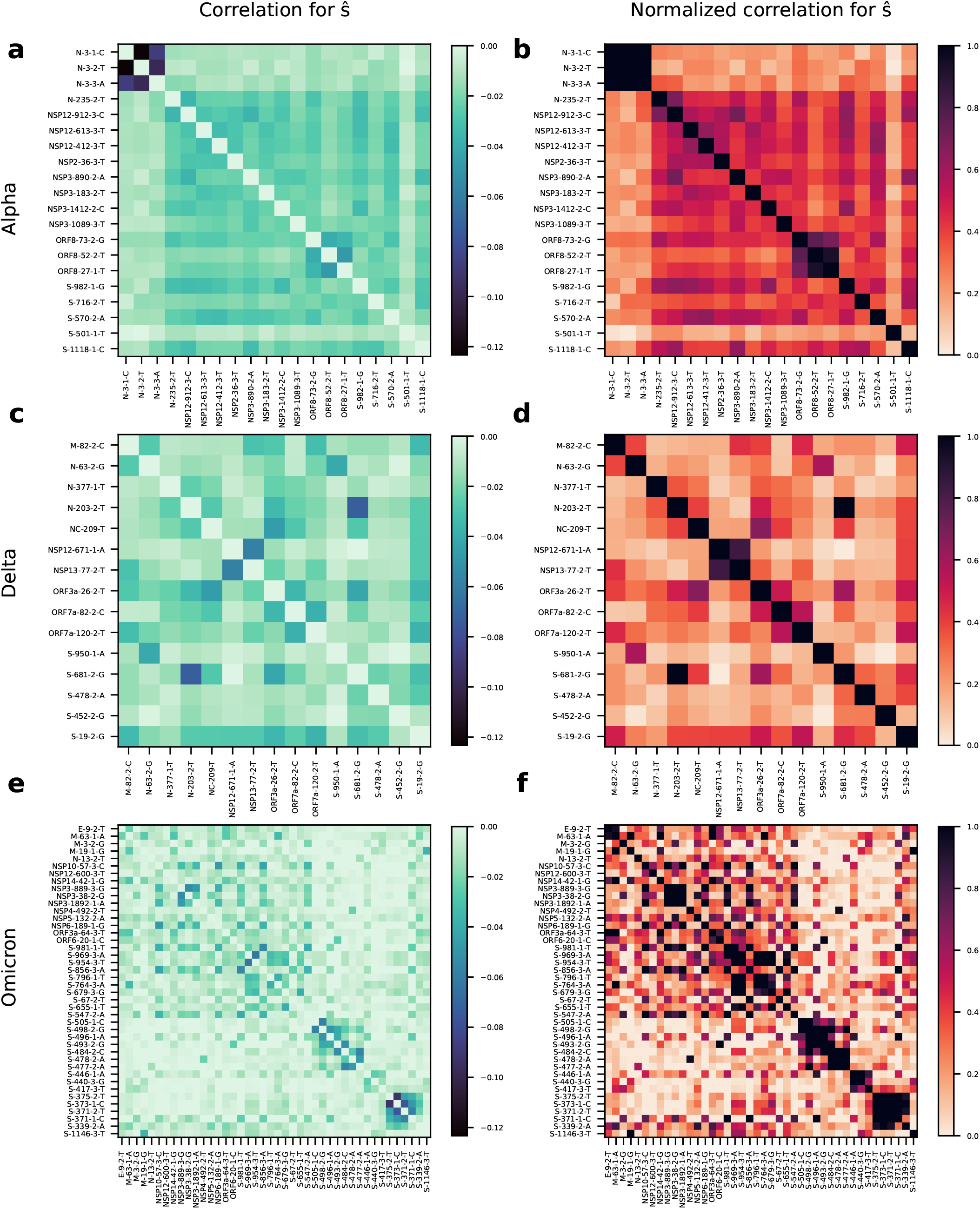
Example correlations between *ŝ* for strongly linked subsets of mutations defining major variants. As discussed in **Supplementary Information**, the covariance of the inferred parameters is given by the matrix in (S5). The correlation matrix of the inferred parameters is easily calculated from this covariance. SNV labels are in the format of xxx-yyy-z-n, where xxx is the protein, yyy is the codon in the protein, z is the index of the nucleotide in the codon, and n is the nucleotide. **a, c, e**, The correlation matrix for SNVs that are strongly linked to one another in Alpha, Delta, and Omicron, respectively. The diagonal elements, all equal to 1 in a correlation matrix, are set to zero for visualization purposes. **b, d, f**, Correlation matrices from **a, c**, and **e**, normalized by the maximum possible correlation for a group of linked SNVs, as discussed in **Supplementary Information**, with the same number of SNVs. The (*i, j*)th element of these matrices represents the percent of linkage between the selection coefficients for SNVs *i* and *j*.

**Supplementary Table 1.** Table of most highly selected amino acid substitutions across the SARS-CoV-2 genome. Error bars were found by taking random sub-samples of 80% of the original regions and re-estimating the selection coefficients. Error bars are the standard deviation of the inferred coefficient for each site over 100 replicates. * represents the cases where phenotypic effect of an amino acid variant has not been reported explicitly in the literature. Instead, it is either based on the function of the encompassing gene, for a mutation to a different amino acid or deletion at the same position. ^#^ all three mutations appear together; RBM = receptor binding motif; RBD = receptor binding domain; NTD= N-terminal domain; FCS = S1/S2 furin cleavage site; HR1 = heptad repeat 1; nAbs = neutralizing antibodies.

| Rank | Protein | Mutation(s)<br>(nt) | Mutation<br>(aa) | Selection<br>(%) | Location | Associated variant(s) | Phenotypic effect |
| --- | --- | --- | --- | --- | --- | --- | --- |
| 1 | S | T23018C/<br>T23019C | F486P | 16.9 ± 0.8 | RBM | XBB.1.5, EG.5.1, BA.2.86, JN.1 | Reduces recognition by neutralizing antibodies<sup>96</sup> |
| 2 | NSP4 | C10029T | T492I | 16.6 ± 0.9 |  | Delta, Lambda, Mu, BA.1 (and subvariants) | Increased viral replication capacity and infectivity, cleavage efficiency of the viral protease, and antibody evasion<sup>97</sup> |
| 3 | NSP6 | T11288-90 | Δ106 | 16.5 ± 0.8 |  | Alpha, Beta, Gamma, Eta, Iota, Lambda, BA.1 (and subvariants) | *Increased transmission by interferon antagonism<sup>98</sup> |
| 4 | S | A23055G | Q498R | 16.2 ± 1.7 | RBM | BA.1 (and subvariants) | Increased ACE2 binding and resistance to nAbs<sup>95,99</sup> |
| 5 | S | A24424T | Q954H | 14.0 ± 1.0 | HR1 | BA.1 (and subvariants) | Increased infectivity in vitro<sup>100</sup> |
| 6 | S | T22942A | N460K | 13.8 ± 0.9 | RBM | XBB, XBB.1.5, EG.5.1, HK.3, BA.2.86, JN.1 | Enhanced neutralization resistance, enhanced spike processing and cell-cell fusion, improves ACE2 binding<sup>101</sup> |
| 7 | S | C23604G | P681R | 13.6 ± 0.9 | FCS | Delta, Kappa, BA.2.86, JN.1 | Enhanced cleavage, fusogenicity, and pathogenicity<sup>102</sup> |
| 8 | S | G22599C | R346T | 13.5 ± 0.5 | RBD | XBB, XBB.1.5, EG.5.1, HK.3 | Evasion of antibody recognition<sup>96</sup> |
| 9 | S | T24469A | N969K | 13.2 ± 0.6 | HR1 | BA.1 (and subvariants) | Improved structural stability<sup>95</sup> |
| 10 | S | T23599A | N679K | 12.6 ± 0.6 | FCS | BA.1 (and subvariants) | *Increased proteolytic activation<sup>95</sup> |
| 11 | S | G22927C | L455F | 12.3 ± 0.4 | RBM | HK.3 (L455S in JN.1) | Enhanced resistance to immune sera<sup>103</sup>, increased ACE2 binding<sup>104</sup> |
| 12 | S | C23525T | H655Y | 11.7 ± 0.9 | FCS | Gamma, BA.1 (and subvariants) | Increased viral replication, spike protein cleavage, and transmission in vivo<sup>105</sup> |
| 13 | N | G28881T | R203M | 11.4 ± 1.2 |  | Delta, Kappa | Enhanced replication, RNA delivery and packaging<sup>106</sup> |
| 14 | S | G22599A | R346K | 10.3 ± 0.5 | RBD | Mu, BA.1, XBB, XBB.1.5, EG.5.1, HK.3 | Reduced neutralization<sup>107</sup> |
| 15 | S | A23063T | N501Y | 10.1 ± 1.1 | RBM | Alpha, Beta, Gamma, Mu, BA.1 (and subvariants) | Increased infection, transmission, ACE2 binding, and resistance to nAbs<sup>95</sup> |
| 16 | S | C21618G | T19R | 9.2 ± 0.7 | NTD | Delta | *Increased resistance to NTD-specific nAbs<sup>108,109</sup> |
| 17 | NSP12 | G15451A | G671S | 9.1 ± 0.6 |  | Delta, XBB, XBB.1.5, EG.5.1, HK.3 |  |
| 18 | S | T22928C | F456L | 8.9 ± 0.9 | RBM | HK.3 | Enhanced resistance to immune sera<sup>103</sup>, increased ACE2 binding<sup>104</sup> |
| 19 | S | C21618T | T19I | 8.7 ± 1.3 | NTD | BA.2 (and subvariants) | *Increased resistance to NTD-specific nAbs<sup>108,109</sup> |
| 20 | S | A22910G | N450D | 8.3 ± 0.6 | RBM | BA.2.86, JN.1 | Increased ACE2 binding<sup>110</sup> |
| 21 | S | C22995G | T478K | 8.2 ± 0.8 | RBM | BA.1 (and subvariants) | T478K enhances ACE2 binding<sup>111</sup>, and enhances neutralization resistance<sup>112</sup> |
| 22 | S | C22916A | L452M | 8.2 ± 0.6 | RBM | BA.2 subvariants; L452W in BA.2.86 and JN.1, L452R in BA.4, BA.5 | Increased RBD expression (stability)<sup>113</sup>, increased resistance to nAbs<sup>114,115</sup>, and increased cell entry<sup>116</sup> |
| 23 | NSP6 | T11296G | F108L | 7.6 ± 1.0 |  |  | *Increased transmission by interferon antagonism<sup>98</sup> |
| 24 | S | Δ21986-88 | Δ142 | 7.6 ± 1.0 | NTD | BA.1 (G142D in BA.2 and subvariants) | *Increased resistance to NTD-specific nAbs<sup>117</sup> |
| 25 | S | C22033A | F157L | 7.4 ± 0.3 | NTD | BA.2.75 (F157S in BA.2.86 and JN.1) | In epitope recognized by neutralizing antibodies<sup>118</sup> |
| 26 | S | A22893G | K444R | 7.3 ± 0.5 | RBM |  | Increased resistance to immune sera<sup>103</sup>, evasion of antibody recognition<sup>96</sup> |
| 27 | N | G28881A | R203K | 7.2 ± 1.0 |  | Alpha, Gamma, Lambda, BA.1 (and subvariants) | Enhanced replication, RNA delivery and packaging<sup>106</sup> |
| 28 | S | T22031- | Δ157 | 7.2 ± 0.5 | NTD | Delta | In epitope recognized by neutralizing antibodies<sup>118</sup> |
| 29 | S | G22577C | G339H | 7.2 ± 0.7 | RBD | XBB.1.5, BA.2.75, EG.5.1, HK.3, BA.2.86, JN.1 | G339D Interferes with T-cell response<sup>119</sup> |
| 30 | S | T23018G | F486V | 7.2 ± 0.5 | RBM | BA.4, BA.5 | Increased ACE2 binding and resistance to nAbs<sup>120,121</sup> |
| 31 | S | T22917A | L452Q | 7.1 ± 0.4 | RBM | Lambda, BA.2.12.1 | Increased RBD expression (stability)<sup>113</sup>, increased resistance to nAbs<sup>114,115</sup>, and increased cell entry<sup>116</sup> |
| 32 | S | T22896A | V445H | 7.0 ± 0.7 | RBM | BA.2.86, JN.1 | Enhanced resistance to immune sera<sup>103</sup>, increased ACE2 binding<sup>104</sup> |
| 33 | S | A22629C | K356T | 7.0 ± 0.4 | RBD | BA.2.86, JN.1 | Neutralization of immune sera<sup>122</sup> |
| 34 | S | C23854A | N764K | 6.9 ± 1.5 |  | BA.1 (and subvariants) | Improved structural stability<sup>123,124</sup> |
| 35 | N | Δ28367-69 | Δ32 | 6.9 ± 1.4 |  | BA.1 (and subvariants) |  |
| 36 | S | C23604A | P681H | 6.7 ± 0.9 | FCS | Alpha, Mu, BA.1 (and subvariants, BA.2.66 and JN.1 have P681R) | Enhanced cleavage<sup>125</sup> and increased resistance to interferon-induced immunity<sup>126</sup>, leading to increased replication and/or transmission |
| 37 | S | G22898A | G446S | 6.6 ± 0.9 | RBM | XBB, XBB.1.5, EG.5.1, HK.3, BA.2.86, JN.1 | enhanced resistance to neutralizing antibodies<sup>101</sup> |
| 38 | NSP6 | T11288A | S106T | 6.6 ± 0.6 |  |  | *Increased transmission by interferon antagonism<sup>98</sup> |
| 39 | N | C28311T | P13L | 6.5 ± 1.4 |  | Lambda, BA.1 (and subvariants) | Escape from a HLA-B*27:05 CD8<sup>+</sup> T cell epitope<sup>127</sup> |
| 40 | S | G23948T | D796Y | 6.4 ± 1.0 |  | BA.1 (and subvariants) | Improved structural stability<sup>95</sup> and antibody evasion<sup>128</sup> |
| 41 | N | C28367T | R32C | 6.4 ± 1.7 |  |  | Alters frustration state of virus and may affect stability, function, and pathogenicity<sup>129</sup> |
| 42 | S | C22674T | S371F | 6.2 ± 0.6 | RBD | BA.2 (and subvariants) | Increased resistance to nAbs<sup>115,130</sup> |
| 43 | S | T22917G | L452R | 6.1 ± 0.6 | RBM | Delta, Kappa, Epsilon, BA.4, BA.5 | Increased RBD expression (stability)<sup>113</sup>, increased resistance to nAbs<sup>114</sup>, and increased cell entry<sup>115</sup> |
| 44 | S | A22893C | K444T | 6.1 ± 0.7 | RBM | BQ.1 | Increased resistance to immune sera<sup>103</sup>, evasion of antibody recognition<sup>96</sup> |
| 45 | S | G23222A | E554K | 6.1 ± 0.5 | RBM | BA.2.86, JN.1 | Escape from monoclonal antibodies<sup>131</sup> |
| 46 | S | C22295A | H245N | 6.0 ± 1.4 | NTD | BA.2.86, JN.1 | Significantly increases ACE2 binding<sup>110</sup> |
| 47 | S | G22775A | D405N | 5.5 ± 1.3 | RBD | BA.2 (and subvariants) | Escapes many neutralizing antibodies<sup>132</sup> |
| 48 | S | C22353A | A264D | 5.3 ± 1.6 | NTD | BA.2.86, JN.1 | Increases ACE2 binding<sup>110</sup> |
| 49 | S | G21987A | G142D | 5.3 ± 0.5 | NTD | BA.2 (and subvariants), BA.4, BA.5 | Increased resistance to NTD-specific nAbs<sup>95,117</sup> |
| 50 | S | G24368T | D936Y | 5.2 ± 1.3 |  |  | Increased infectivity<sup>114</sup> |

## Supplementary Information

### 1. Summary

Here we discuss two main topics. First, we give a detailed introduction of our epidemiological model as well as a derivation of the estimator (1) and an important simplification of it. Second, we describe simulations of an outbreak and show that selection coefficients can be accurately recovered from simulation data even with relatively poor sampling.

### 2. Epidemiological model

#### 2.1 Introduction

In epidemiology, the spread of infection can be modeled as a branching process where each infected individual (also referred to as a case) infects *n* additional individuals ^133^. The distribution of *n* is often taken to be Poisson, but differences in the number of contacts with susceptible individuals, disease course within an individual, and other factors mean that the Poisson rate *λ* is not generally the same for all cases ^134^. Below, we first follow ref. ^134^ to explore families of distributions for the number of new cases per infected individual. Next, we extend these models to consider multiple variants of the pathogen that differ in their spreading efficiency. We seek to characterize how the distribution of pathogen variant frequencies is expected to change over time, and how such data can be used to estimate the relative spreading efficiency of different variants.

#### 2.2 Distributions for the number of infected individuals

As noted above, the basic distribution of the number of new cases *n* caused by one case in a susceptible population is Poisson,

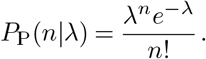

Typically we might take the Poisson rate *λ* to be *R*, the effective reproduction number, which is the expected number of cases directly caused by one case. In that case, the average number of cases following the Poisson distribution is

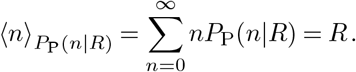

To account for variability in transmission dynamics, the basic Poisson distribution with a single rate *R* can be replaced with a continuous mixture of Poisson distributions, where the rate parameter *λ* follows a gamma distribution,

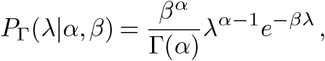

with shape parameter *α* and rate parameter *β*. The average value of *λ* is

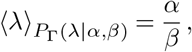

and its variance is

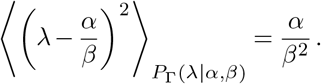

In this context, it is natural to take *α* = *k* and *β* = *k/R*. With these choices, the gamma distribution reads

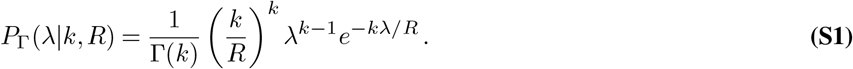

The parameter *k* is a dispersion parameter that determines how long-tailed the distribution is. The mean value of *λ* is always *R*, but when *k* is smaller its variance increases. In the limit that *k*→ ∞, we recover the pure Poisson distribution with rate *λ* = *R*. When *k* = 1, the distribution of the number of cases *n* is geometric,

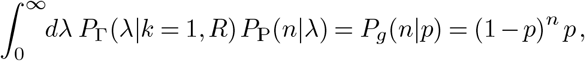

where *p* = 1*/*(1 + *R*). For arbitrary values of *k >* 0, the number of cases follows a negative binomial distribution,

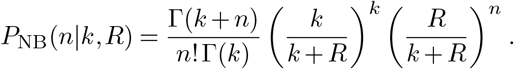

The standard parameters of the negative binomial distribution are *r* and *p*, which are set to *k* and *k/*(*k* + *R*) in our parameterization above.

#### 2.3. Dynamics for variant frequencies

Let us assume that there exist multiple variants of a pathogen, which are distinguished by an index *a*. The number of cases infected with variant *a* is *n*_*a*_. We assume that different variants have slightly different transmission probabilities, so that *R*_*a*_ = *R*(1 + *w*_*a*_), with |*w*_*a*_| *«* 1. The term *w*_*a*_ is analogous to a selection coefficient in population genetics.

##### 2.3.1. Dynamics of multiple cases infected by a single variant

First, let us assume that *n* individuals, each labeled by an index *i*, are all infected by the same variant of a pathogen. How many cases will be generated from these individuals? The number of new cases for all individuals is

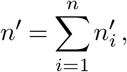

where the numbers of cases 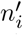 generated by individual *i* follows a negative binomial distribution. Because all individuals are infected by the same variant, the negative binomial parameter *p* = *k/*(*k* + *R*) is the same for each of them. Then, assuming that all of the infection events are independent, it can be shown that the probability distribution for the total number of new cases *n′* also follows a negative binomial distribution with the same value of *p*, and with *r* = *nk* (that is, the new *r* parameter value is the sum of the individual *r* parameter values). Thus, the distribution of *n′* is

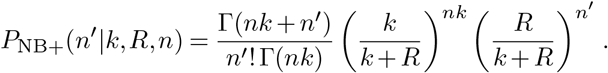

##### 2.3.2. Dynamics for multiple cases infected by multiple variants

Let us extend the previous example to consider *m* variants of a pathogen. At the starting point, the number of individuals infected by a given variant *a* is *n*_*a*_, with *a* ∈ {1,…, *m*}. The fraction of cases infected by variant *a* is

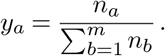

Now, we would like to know how the fraction of individuals infected by each variant is expected to change with each round of infections. In other words, for variant *a*, we would like to compute

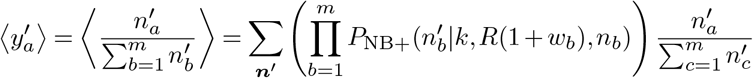

where the outer sum is over all vectors ***n****′* with entries 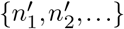, and with 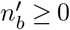 for all *b*. Here, we have assumed that the 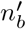’s are independent across *b*.

To proceed, it is convenient to write the negative binomial distributions as mixtures of Poisson distributions (as indicated above), giving

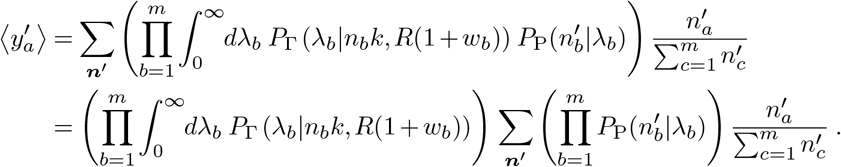

Next, we use the fact that the sum of independent Poisson-distributed random variables is also Poisson with rate parameter equal to the sum of the individual rates, and that the distribution of independent Poisson random variables conditioned on their sum is multinomial, to write

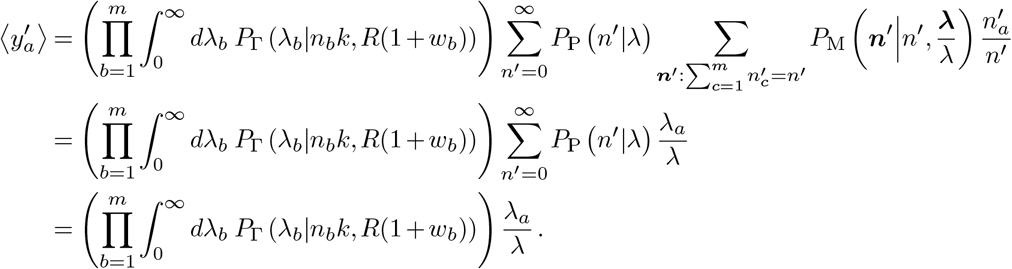

Here ***λ*** is a vector with entries {*λ*_1_, *λ*_2_, …}, and we have also introduced Σ_*a*_ *λ*_*a*_ = *λ*. Note also that the outer sum on the first line is over all vectors ***n′*** whose (non-negative) entries sum to *n′*.

Computing the remaining integrals exactly is challenging, largely because the Gamma distributions have different rate parameters. To address this, next we will expand our expression to first order in the *w*_*a*_, since these are assumed to be small parameters. Referring back to Eq. (S1), the expansion gives

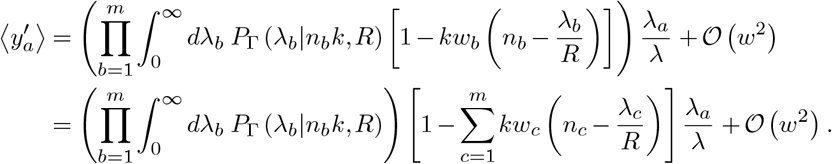

Next we change variables to {*λ, q*_1_ = *λ*_1_*/λ, q*_2_ = *λ*_2_*/λ*,…, *q*_*m*−1_ = *λ*_*m*−1_*/λ*}, because the distribution of the sum of gamma-distributed random variables, *λ*, with the same rate parameter and the ratios of the individual variables to the total (*λ*_*a*_*/λ*) follow independent gamma and Dirichlet distributions ^135^. The *m*th ratio 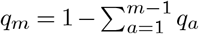 by conservation. By convention we will also set *w*_*m*_ = 0, which can be thought of as normalizing the value of *R* relative to a reference genotype. The transformation then gives

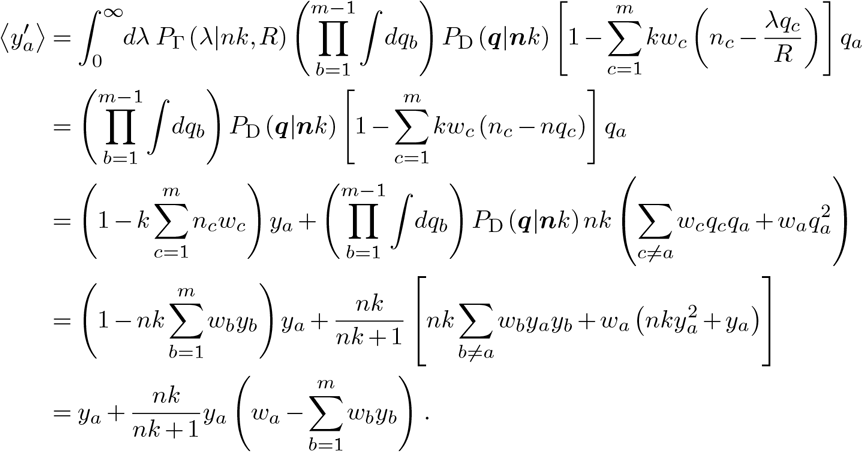

In the expressions above *P*_D_(***q***|***α***) is the Dirichlet distribution, with concentration parameters ***α*** given by ***n****k* in our case. Note that if *w*_*m*_ ≠ 0, the last line should instead read

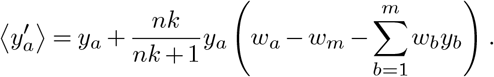

Thus, we obtain (with *w*_*m*_ = 0)

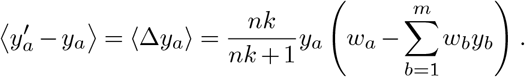

Following a similar approach, we can compute the second moments. First, we consider

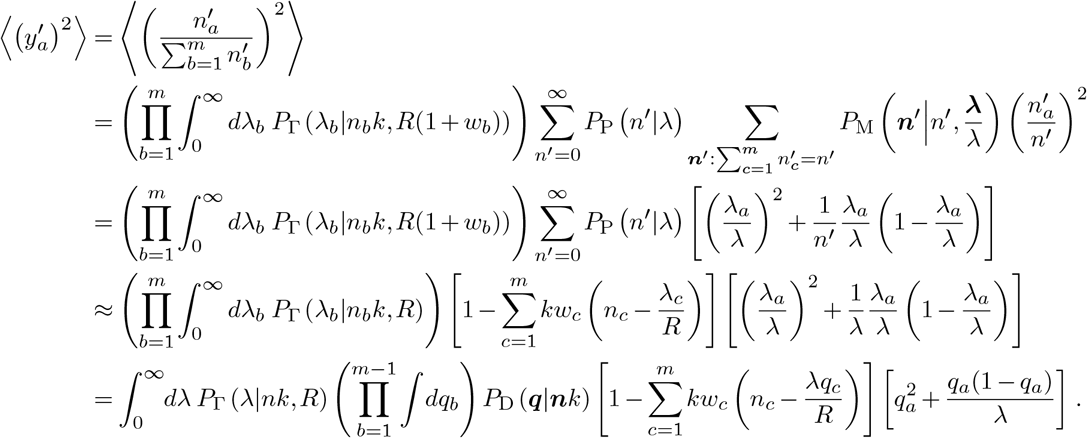

In going from the third to the fourth line above, we have made the approximation that

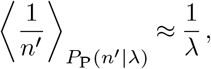

which is valid for *λ* ≳ 1. Similarly,

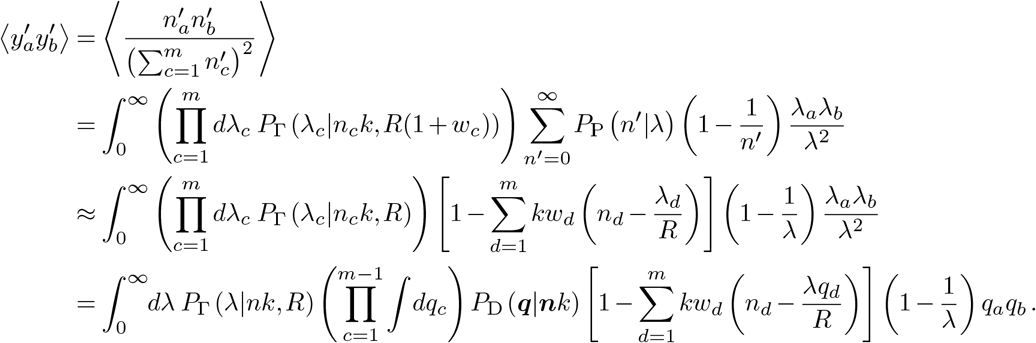

Simplifying the expressions above is tedious but straightforward. The following results are helpful:

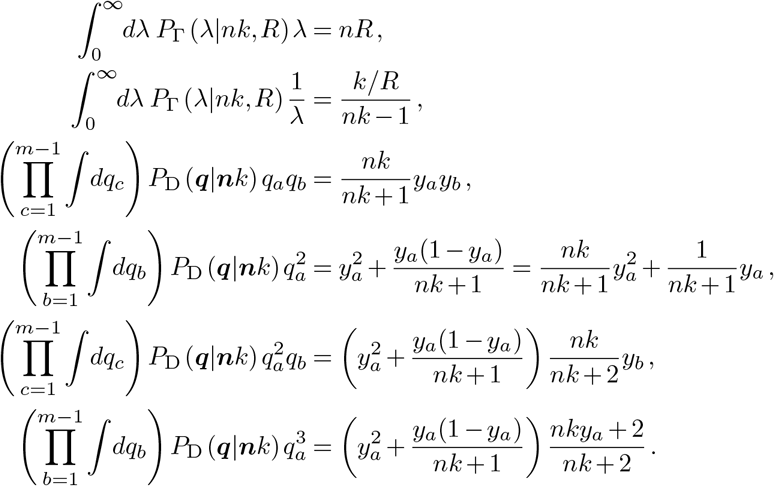

Here we have frequently used *n*_*a*_ = *ny*_*a*_ to simplify expressions.

With the above results, simplifying expressions for the second moments, we finally find

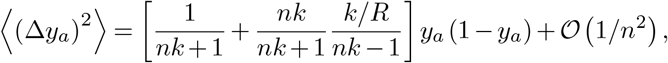

and

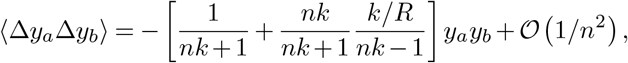

where we have assumed that the *w*_*a*_ are 𝒪 (1*/n*), as in the Wright-Fisher model with weak selection. We have thus found that the first and second moments of frequency changes in our multi-variant epidemiological model have the same frequency dependence as those in the multispecies Wright-Fisher model, but with different scaling. The first moment (‘drift’) is multiplied by a factor of *nk/*(*nk* + 1), and the second moment (‘diffusion’) by

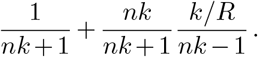

These prefactors match with the Wright-Fisher model exactly when *k*→ ∞ (i.e., a pure Poisson distribution for the number of new cases per infected individual) and *R* = 1.

#### 2.4. Derivation of the selection coefficient estimator

The derivation in this section closely follows that given in ref. ^136^. It is well known that a WF process can be approximated by a continuous-time continuous-frequency diffusion process in the large *n* limit. In the continuous-time limit the time variable *t* has units of *n* generations, with one generation in discrete time taking *τ* = 1*/n* continuous time units. The selection coefficients *w*_*a*_ are assumed to scale with *n* such that 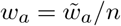, where 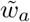 is a parameter independent of the population size *n*. In the limit of large population size, our generalized super-spreading model can, like the WF process, be approximated by a diffusion process, where the transition probability density *ϕ* is the solution to the Fokker-Planck equation

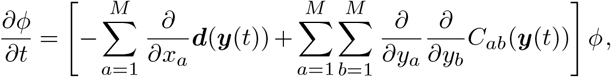

where *M* is the number of distinct genotypes, ***y*** is the genotype frequency vector, ***d*** is the drift vector, and *C* is the diffusion matrix. Here we ignore recombination and mutation, since these are comparatively small and therefore unlikely to significantly affect estimates of changes in viral transmission (though these can be included and the solution remains tractable). The drift and diffusion have entries given by,

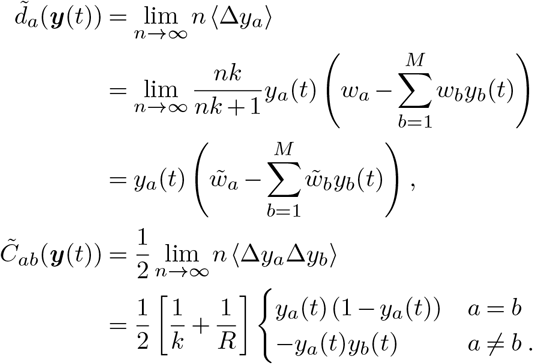

For genotype frequencies observed at times *t* and *t* + *τ* Δ*t* (i.e., over Δ*t* generations), and for small *τ* Δ*t*, the Fokker-Planck equation can be converted into a path integral approximation for the transition probability density (see ref. ^136^ for a rigorous derivation)

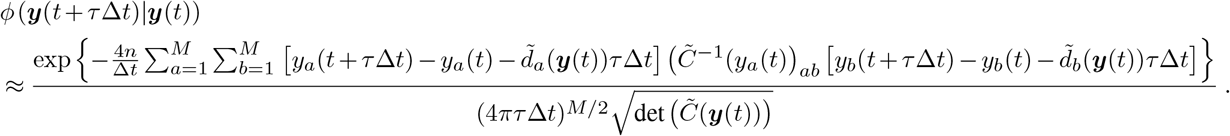

From this result, and recalling *τ* = 1*/n*, the transition probability from time *t*_*m*_ to *t*_*m*+1_ of the original branching process (for large *n/*Δ*t*) can be approximated by

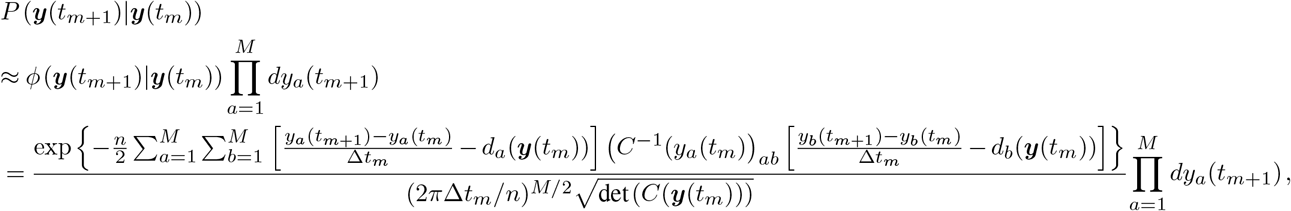

where we write the re-scaled drift vector as 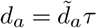, the re-scaled diffusion matrix as 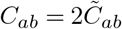, and Δ*t*_*m*_ = *t*_*m*+1_ −*t*_*m*_. Since we aim to infer selection coefficients for the SNVs, it is more convenient to work with the allele frequencies *x*_*i*_ instead of the genotype frequencies *y*_*a*_. The allele frequency at site *i* is given by

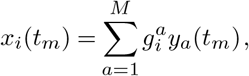

where 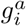 is a 1 if there there is a mutant allele at site *i* on genome *a* and zero if there is not. Similarly, if the selection coefficient for the genotype *a* is *w*_*a*_ and the allele level selection coefficient for allele *j* is *s*_*j*_, then they are related by:

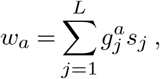

where *L* is the length of the genome.

The allele level drift and diffusion terms will be linear combinations of the genotype level drift and diffusion, just as with the frequencies and the selection coefficients. The drift vector for the allele frequencies can be transformed by

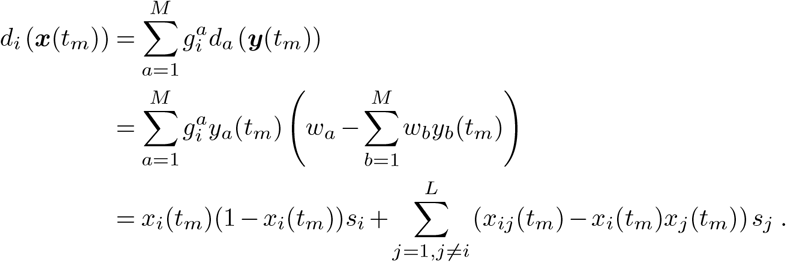

This can be used, along with the transition probability density for genomes, in order to find an approximation for the mutant allele transition probability density:

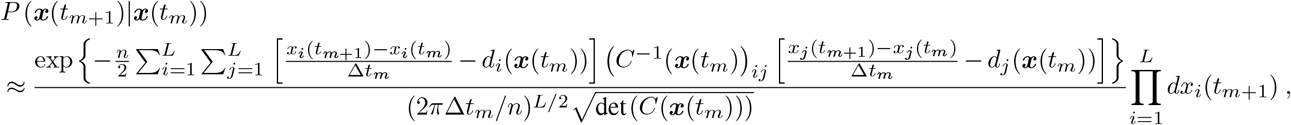

where here the diffusion *C* is derived similarly to the drift ***d*** and has entries

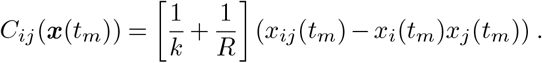

A path integral then gives the probability of observing a trajectory of allele frequencies (***x***(*t*_1_), ***x***(*t*_2_), …, ***x***(*t*_*T* −1_)), and is given by

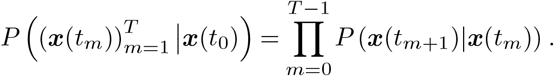

Bayesian analysis can then be used to show that the posterior probability of the selection coefficients ***s*** = (*s*_1_, *s*_2_, …, *s*_*L*_) given an observed frequency path ***x***(*t*_0_), ***x***(*t*_1_), …, ***x***(*t*_*T* −1_) is

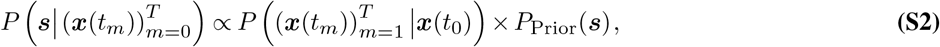

where we use a Gaussian prior distribution with zero mean and adjustable covariance determined by the parameter *γ*, which is the precision.

For the inferred coefficients, we take those that maximize the posterior probability. They can be analytically found by a simple application of the Euler-Lagrange equations to (S2) and are given by

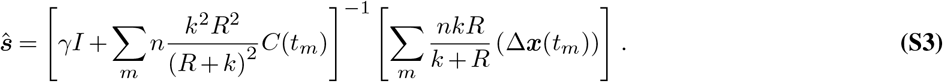

#### 2.5. Extension to multiple regions

In the SARS-CoV-2 pandemic, and in real disease outbreaks in general, there are frequently multiple different outbreaks in different regions that develop largely or entirely independently of one another. In order to find the best estimate for the selection coefficients using the data from multiple regions, the estimator can be generalized to find the maximum a posteriori estimate for the selection coefficients given the time series of allele frequencies in each of the regions. If the probability for a specific path in a specific region *r* is given by 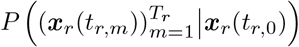, where *x*_*r*_ is the allele frequency vector in region *r*, then the joint probability of the specific paths in all of the regions is simply the product of the individual region probabilities:

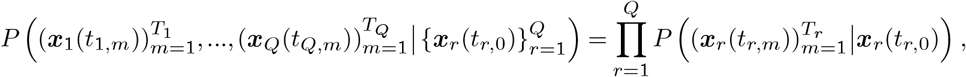

where *Q* is the number of different regions. Since this is a product of exponential functions, the log posterior will be the sum of the exponents and the regularization. This can be maximized with respect to the selection coefficient vector ***s*** as before and leads to the estimator:

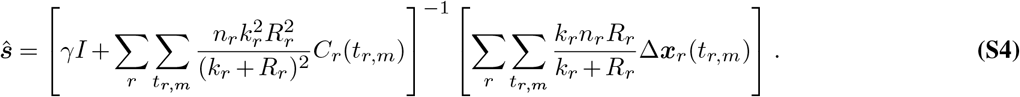

#### 2.6. Simplification of the estimator

In real outbreaks the parameters *k, R*, and *n* are in general time-varying. In our simulations as well, *R* and *n* are time-varying (and *k* can be constant or time-varying). In order to accurately infer the selection coefficients according to Eq. (S3) or Eq. (S4), it would seem that we need to accurately infer the values of *k, R*, and *N* at every point in the time series. In practice, this would be extremely difficult. For general discussion about the effective reproduction number *R* and the basic reproduction number *R*_*t*_ as well as some attempts to infer this, see refs. ^137–141^. In order to get an accurate estimate for *k* it is necessary to have pervasive contact tracing, so that the negative binomial distribution is well sampled, and there are other difficulties in inferring *k* as well ^142–144^. Lastly, it can be difficult to estimate the number of new infections due to multiple factors, including the difference between the population that gets tested and the population that does not, test result inaccuracies, and delays between symptom onset, testing, and reporting.

We propose an alternative that lets us avoid these complications. The prefactor *nkR/*(*R* + *k*), multiplies both the numerator and the denominator. Therefore, the only effect of the prefactor is to weight time points more heavily if the population size, the dispersion parameter, or the basic reproduction number, is larger. This makes sense in theory, because a larger *n* or *k* implies that there is less noise and the trajectories are more deterministic, while a larger *R* means that there are more new infections per generation and thus more data to use to infer the selection coefficients. This does hold with perfect information, that is, if all infected individuals are sampled at every time point. However, in practice, finite sampling is the source of significantly more noise than that due to a time-varying population size or dispersion, so weighting the time points based upon *n, k*, or *R* in fact leads to worse inference than assuming the parameters are constant in time and thus weighting the time points equally. However, in the special and unrealistic case of perfect sampling, using the actual parameters does lead to better inference than using constant parameters (see **Supplementary Fig. 11**). If the time points are weighted equally, then, provided that the regularization *γ* is scaled appropriately (and in general it must be determined by separate means, discussed below), the prefactors in the numerator and denominator cancel, and the estimator is independent of *n, k*, and *R*. Defining *γ′*= *γnkR/*(*k* + *R*) and 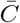 by

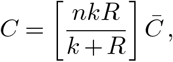

so that

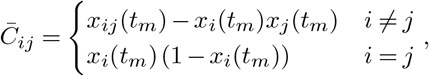

Eqs. (S3) and (S4) for the selection coefficients become, respectively

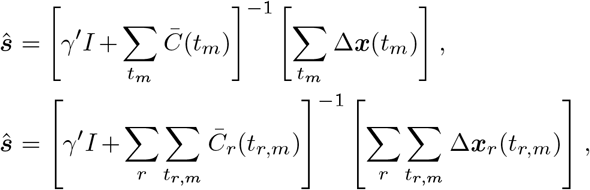

which are the same as the MPL estimators for the Wright-Fisher model except for the absence of a mutation term ^136^.

#### 2.7. Covariance of the inferred selection coefficients

Since the posterior given in (S2) is a Gaussian distribution for the selection coefficients, the covariance matrix of the inferred selection coefficients can be easily found. For any Gaussian distributed random vector ***z***, the inverse of the covariance can be calculated as the second derivative with respect to ***z*** of the negative log of the probability density function. That is, if we define

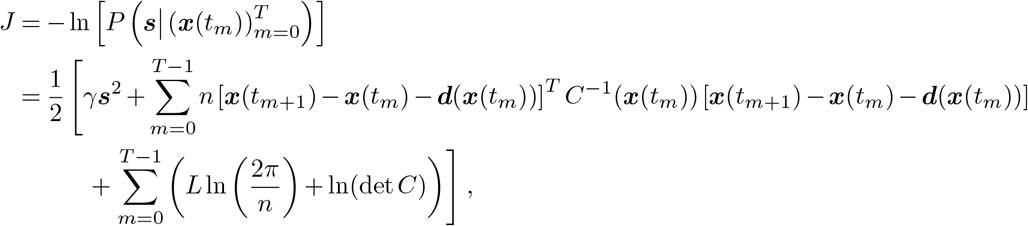

then the inverse of the covariance matrix of the parameters is given by the second derivative of *J* with respect to ***s***. The first derivative of *J* with respect to ***s*** gives

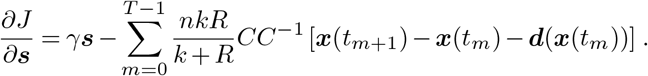

The second derivative, which is the inverse of the covariance of the selection coefficients ***s***, is

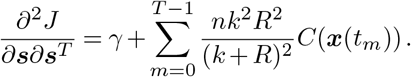

This implies that the covariance of the inferred coefficients is given by

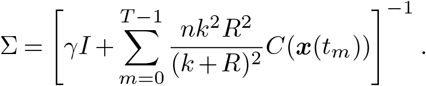

Using the definitions of *γ′* and 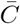 given above, in the case where the parameters *n, k*, and *R* are constant, this reduces to

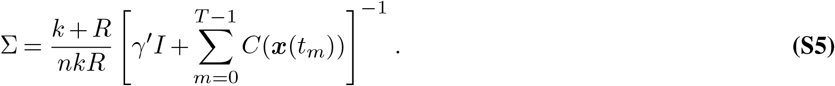

Since (*k* + *R*)*/nkR* is a decreasing function of *k*, this implies that the theoretical covariance decreases as the dispersion *k* becomes larger. **Supplementary Fig. 14a** shows the theoretical uncertainty in the selection coefficients with the largest magnitudes that we infer from SARS-CoV-2 data. Because the theoretical uncertainties do not account for finite sampling, these error bars tend to be fairly small. To obtain more realistic error bars, we also performed bootstrap resampling of the data, where multiple regions were also omitted from the analysis at random (**Supplementary Fig. 14b**.

#### 2.8. Covariance of inferred selection coefficients for a group of fully linked sites

The above analysis can be used to quantify the covariance between inferred coefficients for a group of SNVs that are fully linked, meaning that all of the SNVs in the group appear together on every sequence on which one of the SNVs appear. This is useful because it provides an estimate for the maximum covariance between linked SNVs. An analytical result is presented only for the special case where all of the SNVs under consideration are fully linked, though simulations indicate that the maximum value is not strongly dependent on other SNVs that are partially linked to the main group. The covariance matrix at any time for a group of fully linked SNVs has (*i, j*)th element given by 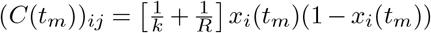 for any (*i, j*), since the frequencies *x*_*i*_(*t*_*m*_) for all of the SNVs are identical. This implies that the second term in (S5) is a matrix with every entry identical. If we define the elements of the matrix

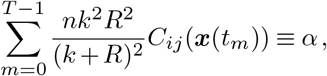

the vector ***u*** as the vector of all 1’s, and use the notation (·)^T^ to denote transpose, then the covariance of the inferred coefficients can be written as

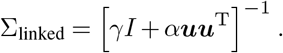

Because of the simplicity of this form of the matrix, the inversion can be carried out explicitly using the Sherman-Morrison formula, which for an *n × n* matrix gives

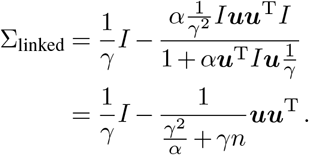

From this the correlation matrix can be easily calculated, and the off-diagonal elements represent the maximum correlation between *n* SNVs that are fully linked to one another. The off diagonal elements of the correlation matrix are given by

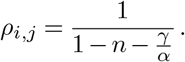

We analyzed sets of strongly linked mutations in the Alpha, Delta, and Omicron variants to test our ability to distinguish the independent selective effects of individual mutations. **Supplementary Figure 15** shows that, while many inferred selection coefficients are naturally correlated, this correlation is far from complete. Only in rare circumstances (e.g., the three nucleotide mutations comprising N:D3L in Alpha) are SNVs so strongly linked that their effects cannot be at least partially disentangled.

### 3. Simulations

We tested the inference using simulations of disease spread. Specifically, we ran super-spreader simulations based on the model described above, which is an analog of the Wright-Fisher model where the sampling distribution for the number of new infections per infected individual is drawn from a negative binomial distribution instead of a pure Poisson distribution.

#### 3.1. Description of simulations

We simulated disease spread as a branching process in which the number of individuals infected per currently infected individual is drawn from a negative binomial distribution whose shape is determined by the basic reproduction number *R*_0_ (or the reproduction number, *R*, in a population that is not totally susceptible) and the dispersion parameter *k*. Because we sample in this way, the population size is not constant. However, if the population size is too small, then the population is extremely likely to die off stochastically, and if the population size is too large, then sampling from the negative binomial becomes too computationally expensive. In order to avoid both of these problems, once the population size is large enough *R* is adaptively adjusted so that the average reproduction number for the entire population will remain near 1, and the population size will oscillate around a fixed value. An explicit time-varying population size can also be used as input, and *R* will be adaptively adjusted to remain near the given curve. Constant values can be used for the dispersion *k* or *k* can vary as a function of time, perhaps representing different degrees of social distancing or lockdown measures at different times. Since different interventions implemented to prevent the spread of disease would likely affect the shape of the distribution of the number of individuals infected by a single infected individual, time-varying values for *k* and *R* can be used to reflect these effects.

#### 3.2. Inference

The simulations are run for a number of generations and genomes are sampled from the population of infected individuals at different times using a multinomial sampling distribution. This sampled time series is then used to infer the selection coefficients using (S3). Alternatively, multiple simulations can be run and the joint inference of the selection coefficients can be made using (S4). We find that, given good enough sampling, a long enough time series, and sampling that occurs at a sufficient number of times, the selection coefficients can be inferred very accurately (**Fig. 1**). The quality of inference is significantly improved if multiple simulations are combined and if mutated sites show up in more than one of the simulations, even under less than ideal sampling conditions. Beneficial coefficients are typically inferred more accurately than deleterious ones, likely because deleterious SNVs frequently die off and therefore there is less data to use for inference.

The inference is robust to shortening the time-series or lowering the number of samples taken per generation, though obviously if either of these conditions is too extreme (or worse, both), the inference starts to break down. The negative effects of a short time-series or poor sampling can be somewhat made up for by using multiple simulations, which is analogous to using data from outbreaks in multiple regions. In addition, the diffusion approximation is only valid in the large *n* limit. However, we tested the inference for small population sizes and found that inference is accurate even if the population of newly infected individuals per serial interval is as low as a few hundred (**Fig. 1**).

